# Critical assessment of variant prioritization methods for rare disease diagnosis within the Rare Genomes Project

**DOI:** 10.1101/2023.08.02.23293212

**Authors:** Sarah L. Stenton, Melanie O’Leary, Gabrielle Lemire, Grace E. VanNoy, Stephanie DiTroia, Vijay S. Ganesh, Emily Groopman, Emily O’Heir, Brian Mangilog, Ikeoluwa Osei-Owusu, Lynn S. Pais, Jillian Serrano, Moriel Singer-Berk, Ben Weisburd, Michael Wilson, Christina Austin-Tse, Marwa Abdelhakim, Azza Althagafi, Giulia Babbi, Riccardo Bellazzi, Samuele Bovo, Maria Giulia Carta, Rita Casadio, Pieter-Jan Coenen, Federica De Paoli, Matteo Floris, Manavalan Gajapathy, Robert Hoehndorf, Julius O.B. Jacobsen, Thomas Joseph, Akash Kamandula, Panagiotis Katsonis, Cyrielle Kint, Olivier Lichtarge, Ivan Limongelli, Yulan Lu, Paolo Magni, Tarun Karthik Kumar Mamidi, Pier Luigi Martelli, Marta Mulargia, Giovanna Nicora, Keith Nykamp, Vikas Pejaver, Yisu Peng, Thi Hong Cam Pham, Maurizio S. Podda, Aditya Rao, Ettore Rizzo, Vangala G Saipradeep, Castrense Savojardo, Peter Schols, Yang Shen, Naveen Sivadasan, Damian Smedley, Dorian Soru, Rajgopal Srinivasan, Yuanfei Sun, Uma Sunderam, Wuwei Tan, Naina Tiwari, Xiao Wang, Yaqiong Wang, Amanda Williams, Elizabeth A. Worthey, Rujie Yin, Yuning You, Daniel Zeiberg, Susanna Zucca, Constantina Bakolitsa, Steven E. Brenner, Stephanie M Fullerton, Predrag Radivojac, Heidi L. Rehm, Anne O’Donnell-Luria

## Abstract

**Background:** A major obstacle faced by rare disease families is obtaining a genetic diagnosis. The average “diagnostic odyssey” lasts over five years, and causal variants are identified in under 50%. The Rare Genomes Project (RGP) is a direct-to-participant research study on the utility of genome sequencing (GS) for diagnosis and gene discovery. Families are consented for sharing of sequence and phenotype data with researchers, allowing development of a Critical Assessment of Genome Interpretation (CAGI) community challenge, placing variant prioritization models head-to-head in a real-life clinical diagnostic setting.

**Methods:** Predictors were provided a dataset of phenotype terms and variant calls from GS of 175 RGP individuals (65 families), including 35 solved training set families, with causal variants specified, and 30 test set families (14 solved, 16 unsolved). The challenge tasked teams with identifying the causal variants in as many test set families as possible. Ranked variant predictions were submitted with estimated probability of causal relationship (EPCR) values. Model performance was determined by two metrics, a weighted score based on rank position of true positive causal variants and maximum F-measure, based on precision and recall of causal variants across EPCR thresholds.

**Results:** Sixteen teams submitted predictions from 52 models, some with manual review incorporated. Top performing teams recalled the causal variants in up to 13 of 14 solved families by prioritizing high quality variant calls that were rare, predicted deleterious, segregating correctly, and consistent with reported phenotype. In unsolved families, newly discovered diagnostic variants were returned to two families following confirmatory RNA sequencing, and two prioritized novel disease gene candidates were entered into Matchmaker Exchange. In one example, RNA sequencing demonstrated aberrant splicing due to a deep intronic indel in *ASNS*, identified in *trans* with a frameshift variant, in an unsolved proband with phenotype overlap with asparagine synthetase deficiency.

**Conclusions:** By objective assessment of variant predictions, we provide insights into current state-of-the-art algorithms and platforms for genome sequencing analysis for rare disease diagnosis and explore areas for future optimization. Identification of diagnostic variants in unsolved families promotes synergy between researchers with clinical and computational expertise as a means of advancing the field of clinical genome interpretation.

## INTRODUCTION

Genome sequencing is increasingly becoming a standard genetic test for rare disease diagnosis and research (1,2), capturing variants in both the coding and non-coding genomic space, and resulting in approximately 75,000 rare variants at ≤1% population allele frequency, per individual, for clinical consideration (3). Therefore, the reported diagnostic gap where >50% of rare disease patients remain undiagnosed becomes more of a question of our capability to interpret, rather than to capture, variation (4,5). The current standards for classification of variant pathogenicity have been defined by the American College of Medical Genetics and Genomics and the Association for Molecular Pathology (ACMG/AMP) and refined by ClinGen (https://clinicalgenome.org/working-groups/sequence-variant-interpretation/) (6–8). They require in-depth assessment of variants to reach pathogenic (P) or likely pathogenic (LP) designation. This well-recognized analytical obstacle to diagnosis is eased by the prioritization of a manageable number of variants for clinical review, and underscores the need to develop computational methods able to integrate variant evidence, such as population allele frequency and *in silico* prediction of deleteriousness, in the context of phenotype and segregation of the variant(s) in the family (9).

CAGI challenges provide a framework to compare genome interpretation methods independently and objectively (10). Predictors are given data and make blind predictions on unpublished datasets which are independently assessed. To understand the state of the art and to encourage innovation for rare disease variant prioritization in a real-life diagnostic setting, we developed the CAGI RGP challenge, implemented in the 6^th^ edition of CAGI assessments (CAGI6). The RGP (raregenomes.org/) study generates and analyzes research genome sequencing data from a diverse range of families seeking a molecular diagnosis for a rare disease. When variants of clear or potential diagnostic relevance are identified, they are clinically validated and returned to participants via their local physicians. For the CAGI6 challenge, we provided predictors with variants from genome sequencing and phenotype data standardized as Human Phenotype Ontology (HPO) terms (11) from a subset of solved and unsolved RGP families. The predictors in the challenge were tasked with identifying the causal variant(s) in as many families and at the highest rank as possible.

Here, we report on the format, assessment, and outcome of the challenge, including lessons learnt from exploration of differences in performance across prediction strategies and provision of method reports from participating teams.

## METHODS

### Sequencing, variant calling, and analysis by the RGP team

Genomic data were obtained by sequencing DNA purified from blood. Sequencing was performed by the Broad Institute Genomics Platform on an Illumina sequencer to 30x depth on average. Raw sequence reads were reassembled against the GRCh38 reference genome. Variants were called with GATK version 4.1.8.0 (12) in the form of single nucleotide variants (SNVs) and small insertions/deletions (indels). All data were analyzed by expert RGP variant analysts using *seqr*, an open-source, web-based genomic analysis tool for family-based monogenic disease analysis (seqr.broadinstitute.org/) (13). Structural variants (SVs) were not included in this challenge, but have been analyzed by the RGP team independent from the CAGI challenge.

### Challenge datasets

Two datasets were provided for the CAGI6-RGP challenge, a training set and a test set. For each, a joint variant call file (VCF) was provided to the CAGI6 organizers for use in the challenge. In addition to the genomic data, clinical phenotype descriptions from patient provided information and review of medical records by a genetic counselor or medical geneticist were provided in HPO nomenclature. The diversity of phenotypes represented the range of clinical presentations routinely seen in patients referred for genetic testing. The family structure and affected status of each sequenced individual were provided, identifying the proband, sibling, mother, and father, as applicable.

For training and contextual purposes, genome sequencing and HPO data from 35 solved RGP families were provided along with the causal variant(s) identified by the RGP team. Ancestry was not provided but was imputed for the probands using the principal component analysis and random forest model used for the Genome Aggregation Database (gnomAD) (3). Overall, the training set consisted of six proband-only families, three duos (proband and one biological parent), and 26 trios (proband and both biological parents). The majority of diagnoses were *de novo* (n=21), or recessive (n=8) and the majority of responsible variants had been reported in the ClinVar database as P and/or LP (14) (https://www.ncbi.nlm.nih.gov/clinvar/) at the time the challenge was announced (May 3, 2021) (**Supplemental Table 1**).

For test purposes, the RGP team selected 30 families for inclusion in the challenge. Fourteen were solved and 16 were unsolved. The solved families in the test set were selected more stringently than for the training set, according to the following criteria: i) the responsible gene has an established Mendelian disease-association as per the Online Mendelian Inheritance in Man database (OMIM, https://www.omim.org/) and/or published literature at the time the challenge was announced, ii) the responsible variant(s) must not have been reported as P/LP in the ClinVar database or listed in/reported as a disease mutation (DM) in the HGMD Professional database (15) (https://www.hgmd.cf.ac.uk/ac/index.php) at the time the challenge was announced (May 3, 2021), and iii) the variant(s) were classified as P, LP, or variant of uncertain significance (VUS) with evidence that is close to LP according to the ACMG/AMP guidelines (6). The causal variants in all 14 solved families had been discussed by the RGP multi-disciplinary team of physicians, genetic counselors, analysts, and molecular geneticists, and had been returned to the family via a local clinician following confirmation in a CLIA certified laboratory. The local clinicians concurred that the variants are diagnostic. The submission of these variants from RGP participants to ClinVar was intentionally delayed for the duration of the challenge. **Supplemental Table 2** displays the answer key for the 30 families in the test set. Overall, the test set consisted of two proband-only families, three duos, 23 trios (proband and both biological parents), and two quads (proband, affected biological sibling, and both biological parents). All of the 16 unsolved families were trios, with the exception of one quad.

A summary of the core features of the families and diagnostic variants in the training and test sets is depicted in **Figure 1**.

**Figure 1.**
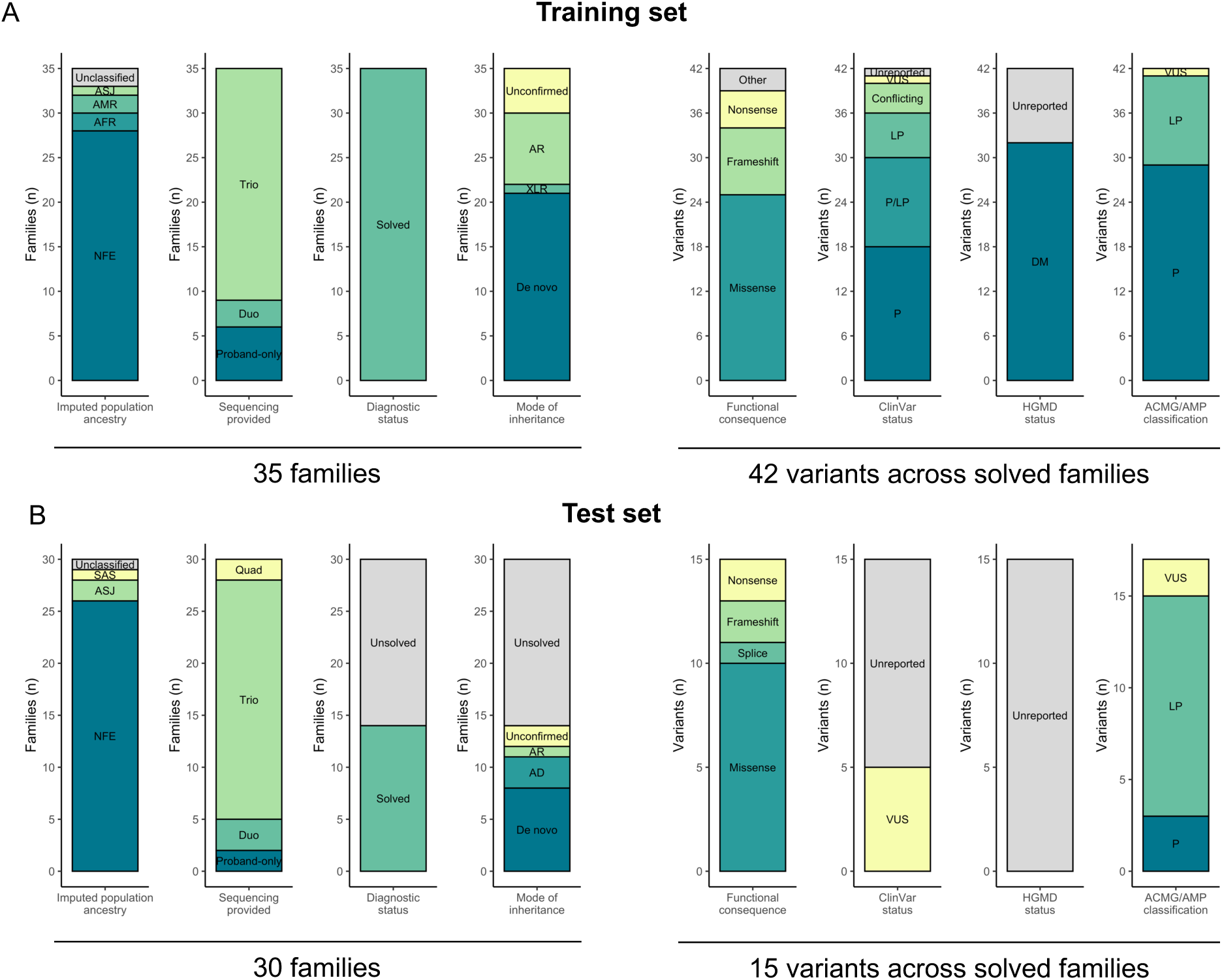
CAGI6 RGP challenge overview of selected families. Summary of the 35 training set families (all solved) and 30 test set families (14 solved, 16 unsolved). Imputed population ancestry, the amount of familial sequencing data provided (proband-only, duo, trio, or quad), diagnostic status, and mode of inheritance of the causal variant(s) is displayed by family. For all returnable diagnostic variants in the solved families in each set, the functional consequence according to the Variant Effect Predictor (VEP), ClinVar and HGMD reporting status at the time of announcement of the challenge (May 3, 2021), and ACMG/AMP classification are displayed by variant. NFE, Non-Finnish European; AFR, African/African American; AMR, Admixed American; ASJ, Ashkenazi Jewish; SAS, South Asian; AD, autosomal dominant; XLR, X-linked recessive; AR, autosomal recessive; P, pathogenic; LP, likely pathogenic; VUS, variant of uncertain significance; DM, disease mutation.

### Challenge format

The challenge was publicly released on May 3, 2021, and open for submissions on June 8, 2021. The submission deadline was October 11, 2021. Participating teams were tasked to provide a genetic diagnosis to as many probands from the 30 families in the test set as possible by submitting predictions for each proband’s causal variant(s). The 14 solved families were included in the challenge to evaluate the performance of each model in prioritizing the causal variants (true positives). The unsolved families were included with the goal of the participating teams identifying novel, potentially causal, variants for further clinical and experimental assessment followed, where possible, by return to the families. The number of solved and unsolved families was not disclosed in the challenge description to allow the participating teams to perform the task in a manner that reflects analysis in the clinical setting. Teams were able to submit up to 100 variant predictions per proband, ranked by causal likelihood, from a maximum of six different models. The submission format, a tab-delimited text file, accepted both single (one variant per line) and proposed biallelic (two variants per line), compound heterozygous predictions. For each variant, teams were required to provide an EPCR value for the submitted variant(s) being causal on a scale of 0 to 1, with 1 indicating highest certainty. An example submission file and a validation script were provided. Predictors were informed that assessors will review how often these were the top variant(s) returned (e.g., in the top 5, 10, 20, 50, or 100 variants) but not further informed about details of the assessment metrics.

### Assessment of model performance across solved families

Formatting errors in the submission files were corrected, and redundant, duplicate, and incomplete submissions were removed. Causal variant predictions for each solved proband were assessed by an independent assessor (author S.L.S). The assessor was blinded to the identity and methods of the participating teams throughout assessment, and the identities of the participating teams were only revealed once the analysis was completed. The following two numeric metrics were considered:

(i) **Mean rank points:** The mean of a weighted point allocation system based on the rank position of the true positive causal variant(s) in the solved probands within the top five (100 points), top 10 (50 points), top 20 (25 points), top 50 (10 points), or top 100 (5 points) variant predictions per proband. Model performance was subsequently ranked by the mean points awarded per proband.
(ii) **F-max:** The F-measure, a harmonic mean between the precision and recall for causal variant prediction in the solved probands, was calculated for all unique EPCR values for each model. The maximum F-measure (F-max) (16), corresponding EPCR threshold, and mean number of predictions submitted per proband at and above this EPCR threshold were subsequently defined for each model and model performance was ranked by the resultant F-max value.

For both numeric metrics, a bootstrapped standard error (SE) (17) was calculated over 1,000 bootstrapped samples from the probands of the 14 solved families in the test set only.

As described, the causal variants in the answer key had been formally classified as P, LP, or VUS leaning towards LP according to the ACMG/AMP guidelines; however, for the purpose of matching the teams’ predictions to the answer key, all variants were treated equivalently. In the case that a correct causal variant was submitted in combination with a second non-causal variant in a proposed biallelic, recessive prediction, the prediction was considered incorrect. For P27, a proband from a family where both the proband and the affected sibling had inherited two paternal variants in *cis*, where it is unknown if both or only one of the variants is required and both variants were considered equally likely to be causal by the RGP team (**Supplemental Table 2**), the highest-ranked variant prediction for either one of the two variants by the respective model was retained and the other was removed from the analysis.

### Assessment of novel putative causal variants across solved and unsolved families

Following assessment of model performance, predictions from top performing models that i) deviated from the answer key in the solved probands and ii) were submitted for the unsolved probands, were critically evaluated in the rare disease genomics web-based analysis tool *seqr* (seqr.broadinstitute.org/) (13). Putative causal variants were discussed by the RGP team and, where possible, were pursued by: i) functional validation of two cases by RNA sequencing, ii) SV analysis in a separate call set generated by the GATK-SV pipeline (18) and manually reviewed in the Integrative Genome Viewer (IGV) (19) to search for a second biallelic, compound heterozygous variant in the case of recessive disease genes, and iii) submission to the Matchmaker Exchange (matchmakerexchange.org/) via *seqr* in the case of candidate novel disease-genes.

### Ethical considerations

The challenge data were derived from patients with rare, suspected monogenic, conditions and their close biological relatives, and included families who are medically underserved (20). Identification of putative causal variants, i.e., causal with respect to the clinical phenotype under investigation, may, if confirmed, be important for tailoring clinical interventions and obtaining social services. We did not actively search for variants unrelated to the rare condition in the family but the consent allows us to optionally provide clinical confirmation of secondary findings if they are incidentally discovered. For the purpose of this challenge, participating teams were told that pathogenic variants unrelated to the proband’s phenotype, such as might be identified as secondary or incidental variants in this challenge (21), should not be returned. All RGP participants have a consent video or phone call with a trained research coordinator to review the study protocol which includes provisions for sharing de-identified data and provide signed informed consent (Mass General Brigham IRB protocol 2016P001422). An institutionally signed (Broad-Northeastern) data transfer agreement was executed. We applied a registered access model (22) where all CAGI6 challenge predictors were required to sign and adhere to the CAGI Data Use Agreement (genomeinterpretation.org/data-use-agreement.html) but institutional signatures were not required.

## RESULTS

### Summary of submissions

Sixteen teams participated in the challenge, submitting predictions from a total of 52 models (median three models per team, range 1-6). Five teams elected to remain anonymous in the reporting, including one team (Team 6) that discovered a bug in their code during assessment and subsequently withdrew from the challenge. Between 0-100 variant predictions (single or proposed biallelic) were submitted per proband (range 0-100, median 100, mean 65). EPCR values ranged from 0-1 (median 0.32, mean 0.38) (**Supplemental Figure 1**). Ninety percent of predictions were single variants and 10% were proposed biallelic, compound heterozygous variants, including entries from five teams that only predicted single variants. Over half (53%) of all variant predictions were in established OMIM disease-associated genes, including entries from six teams limiting analysis to a subset of genes already implicated in disease, though the gene sets differed between groups. Eighty-four percent of predictions were in the coding sequence or direct splice region, as defined by VEP (i.e., within 1-3 bases of the exon, 3-8 bases of the intron, or in the splice polypyrimidine tract). Concordance between models for the top five ranked predictions per proband across all 30 families in the test set ranged from 0-1 (mean 0.09, standard deviation [SD] 0.15) and was significant only between different models from the same team (**Supplemental Figure 2**).

### Summary of numeric assessment of model performance and methodology

Overall, model performance was highly variable (**Figure 2A**). All causal variants in the answer key were predicted within the first five rank positions by at least one model (**Table 1**). Our selected numeric assessment metrics for each submitted model are displayed in **Table 2** and are depicted in **Figure 2B**. One of the top performing models from Team 9 (Invitae Moon) was able to call 13 of the 14 causal variants within the top five rank positions, followed by Team 12 (Lichtarge) with 12, Team 11 (enGenome) and Team 14 (TCS) tied with 10, and Team 5 (Exomiser) with 9 (**Table 2**). Here, we provide a summary of the numeric assessment of model performance and methodology for the five top performing teams. More detailed methods descriptions are provided for in total 11 of the 16 participating teams in the **Supplemental Material**.

**Figure 2.**
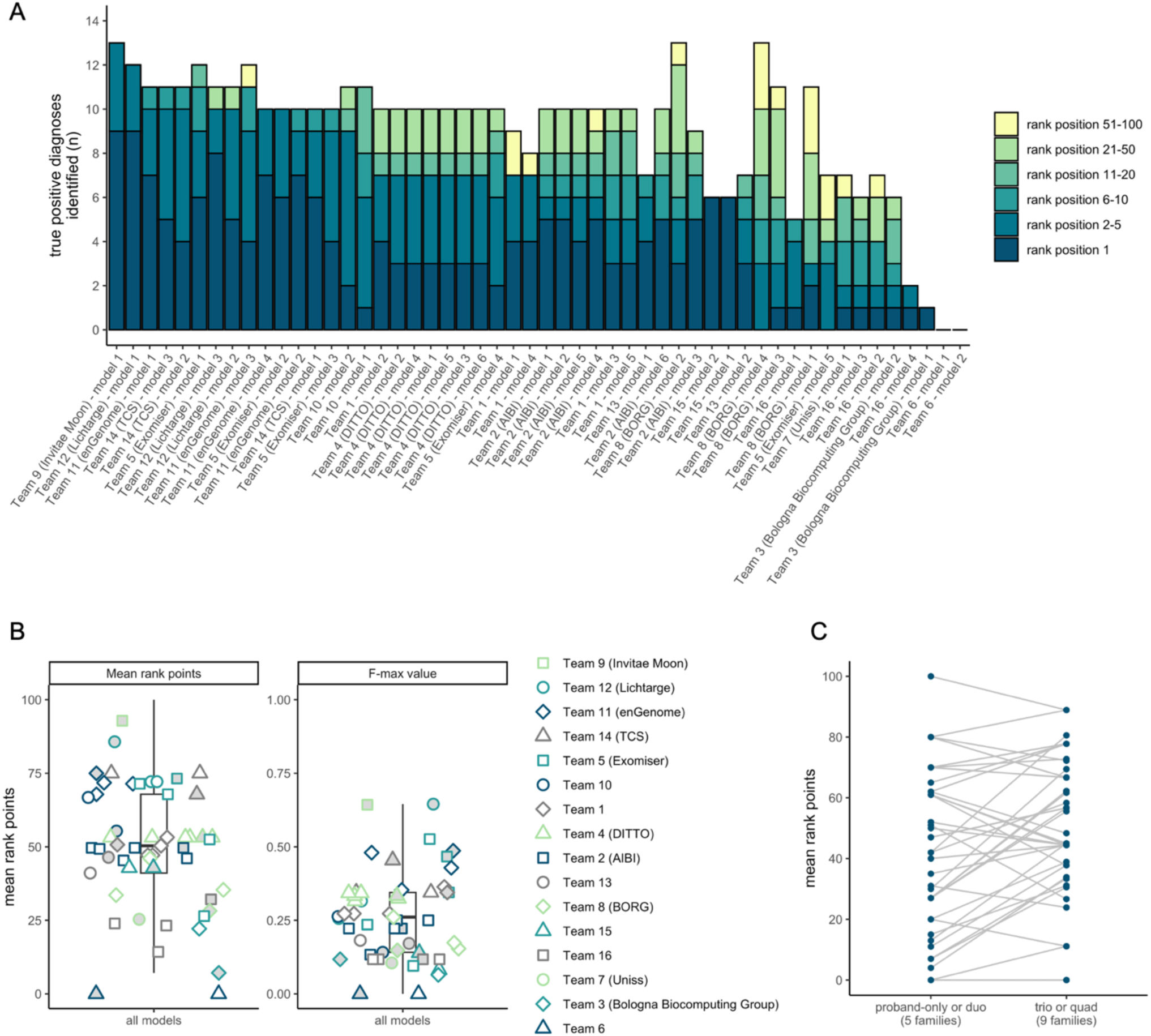
Results of assessment using the 14 solved families (true positives). **A.** Number of true positive diagnoses (y-axis) Identified per model (x-axis) colored by the rank position of the causal variants in the 14 solved probands. Models are ordered by their performance according to the mean rank points metric (**Table 2**). Team names are provided except for teams that elected to remain anonymous. **B.** Results of the mean rank points and F-max value numeric assessment metrics by team and model. Model 1, the primary model, for each team is indicated by the grey fill. **C**, Performance of models, according to the mean rank points awarded, comparing families with proband-only or duo data (i.e., an incomplete trio/quad) versus trio or quad data (i.e., a complete trio/quad).

**Table 1.** Detection of causal variants in the test set summarized across all 52 submitted models.

| Proband | Gene symbol | Inheritance pattern | Variant functional consequence | Sequencing provided | ACMG/AMP classification | Number of models with the causal variant(s) at rank 1 | Number of models with the causal variant(s) at rank 1-5 | Number of models with the causal variant(s) at rank 1-10 | Number of models with the causal variant(s) at rank 1-20 | Number of models with the causal variant(s) at rank 1-50 | Number of models with the causal variant(s) at rank 1-100 |
| --- | --- | --- | --- | --- | --- | --- | --- | --- | --- | --- | --- |
| P28 | *FGFR2* | *de novo* | missense | trio, unaffected parents | P | 36 | **46** | 46 | 47 | 47 | 47 |
| P11 | *TPM2* | dominant | missense | proband-only | VUS | 17 | **40** | 41 | 44 | 47 | 47 |
| P4 | *NALCN* | *de novo* | missense | trio, unaffected parents | LP | 27 | **37** | 39 | 40 | 44 | 46 |
| P7 | *EHMT1* | *de novo* | frameshift | trio, unaffected parents | LP | 27 | **34** | 36 | 37 | 38 | 40 |
| P2 | *FAM111B* | dominant | missense | proband-only | LP | 15 | **27** | 33 | 35 | 41 | 41 |
| P21 | *BICC1* | *de novo* | missense | trio, unaffected parents | LP | 3 | **21** | 25 | 28 | 33 | 36 |
| P24 | *KMT2D* | *de novo* | missense | trio, unaffected parents | LP | 9 | **19** | 28 | 30 | 34 | 34 |
| P22 | *GRIN2A* | unknown | nonsense | duo, unaffected mother | P | 6 | **19** | 23 | 32 | 34 | 34 |
| P23 | *GNAI1* | *de novo* | missense | trio, unaffected parents | LP | 14 | **19** | 19 | 21 | 29 | 31 |
| P16 | *DLG4* | *de novo* | frameshift | trio, unaffected parents | LP | 10 | **18** | 22 | 24 | 25 | 26 |
| P27 | *TUBB8* | dominant | missense | quad, affected sibling | VUS | 11 | **18** | 20 | 22 | 23 | 23 |
| P19 | *CLTC* | *de novo* | splice acceptor | trio, unaffected parents | LP | 8 | **13** | 16 | 17 | 18 | 18 |
| P5 | *PI4KA* | recessive | nonsense; missense | duo, unaffected father | P; LP | 6 | **9** | 12 | 12 | 15 | 18 |
| P6 | *KCND2* | unknown | missense | duo, unaffected father | LP | 0 | **3** | 5 | 12 | 22 | 26 |

The 14 solved (true positive) cases are displayed with the causal gene, inheritance pattern, functional consequence of the causal variant(s), amount of familial sequencing provided in the challenge, and the ACMG/AMP classification of the variant(s). The number of models, out of 52, ranking the variant at position 1, 1-5, 1-10, 1-20, 1-50, and 1-100 are depicted. The probands are displayed in decreasing order by the number of causal variants submitted at rank position 1-5 by the models (emboldened), considered reasonable performance for a prediction metric. P, pathogenic; LP, likely pathogenic; VUS, variant of uncertain significance.

**Table 2.**
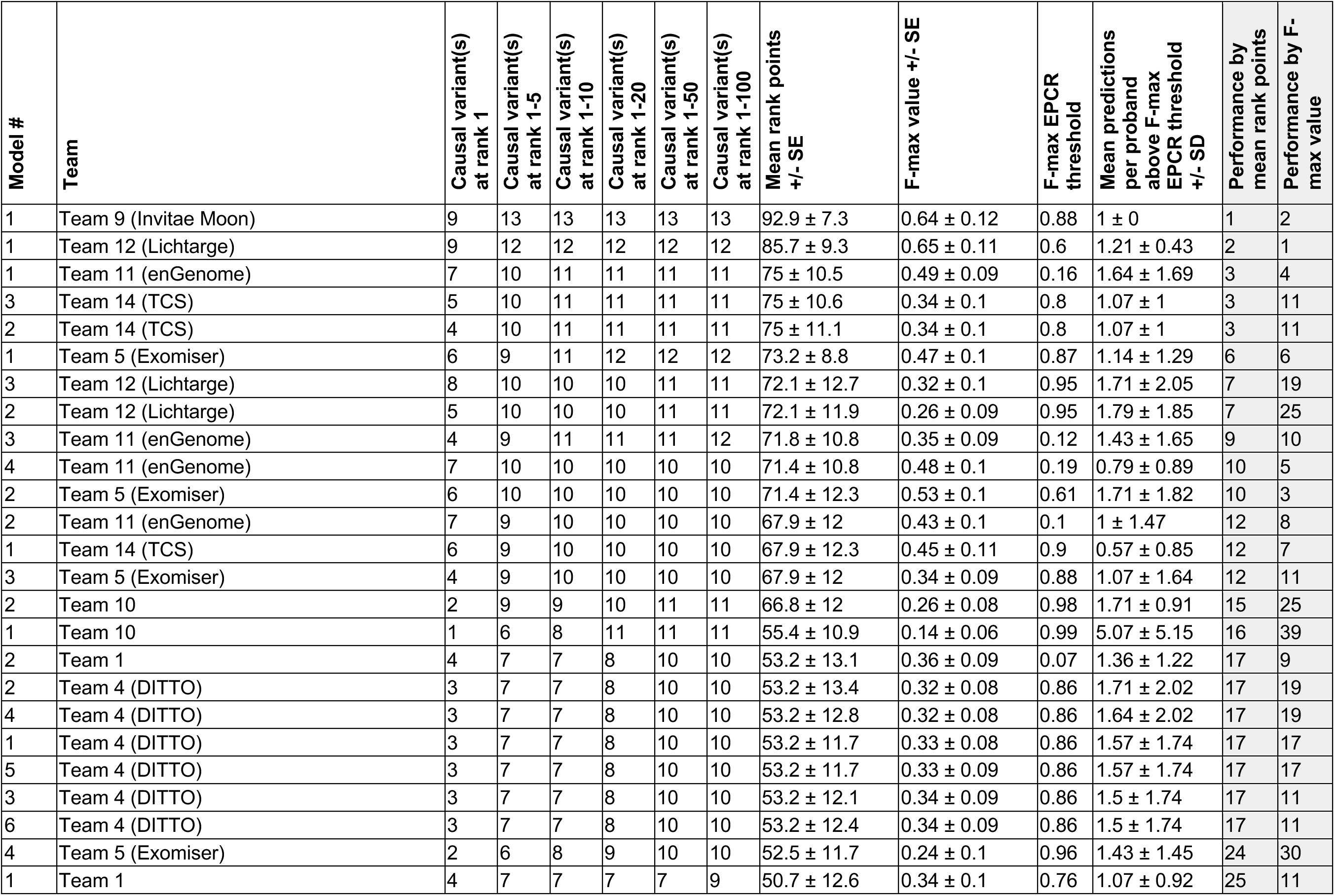
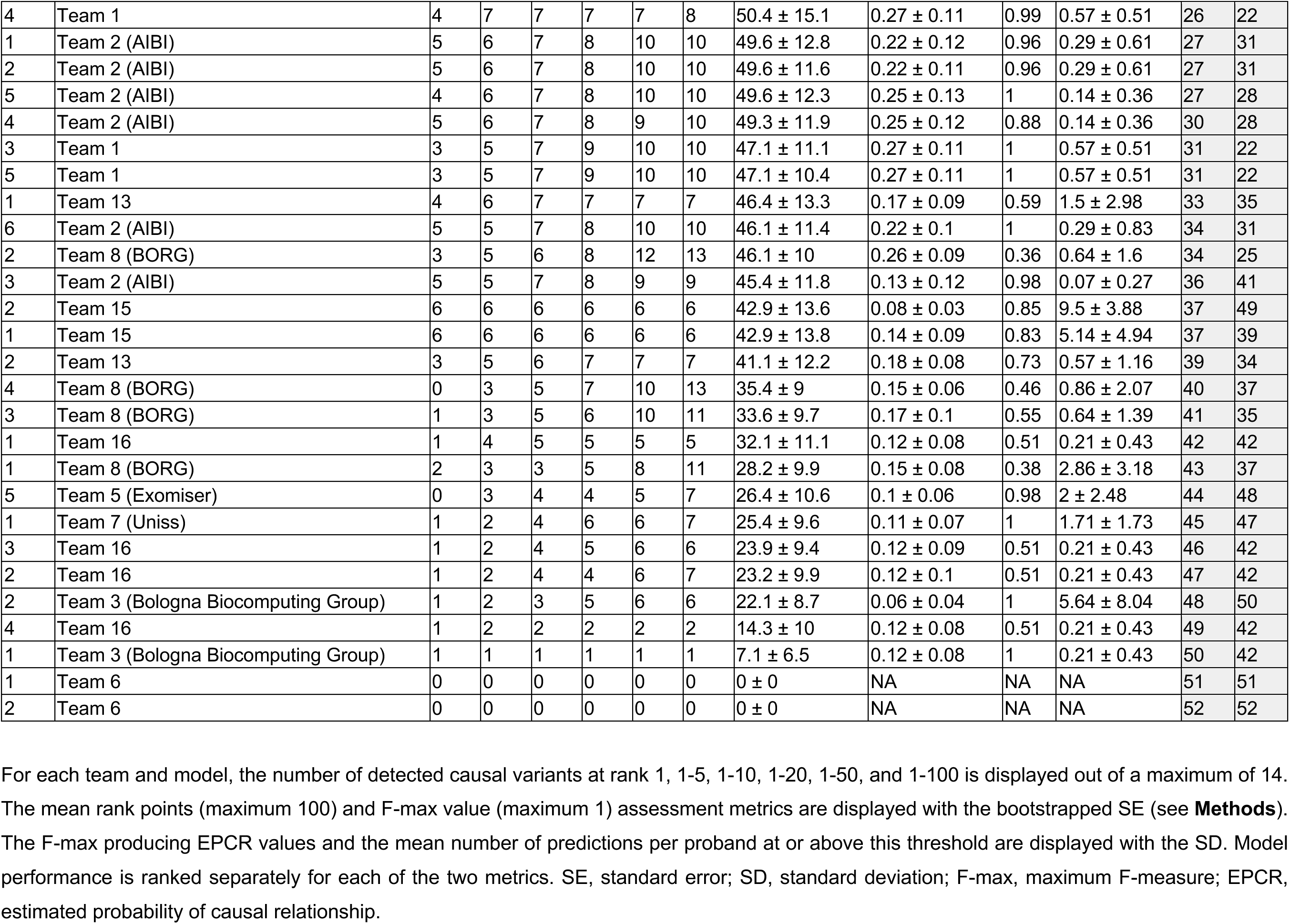
Numeric assessment metrics for all models.

| Model # | Team | Causal variant(s)<br>at rank 1 | Causal variant(s)<br>at rank 1-5 | Causal variant(s)<br>at rank 1-10 | Causal variant(s)<br>at rank 1-20 | Causal variant(s)<br>at rank 1-50 | Causal variant(s)<br>at rank 1-100 | Mean rank points<br>+/- SE | F-max value +/- SE | F-max EPCR<br>threshold | Mean predictions<br>per proband<br>above F-max<br>EPCR threshold<br>+/- SD | Performance by<br>mean rank points | Performance by F-max value |
| --- | --- | --- | --- | --- | --- | --- | --- | --- | --- | --- | --- | --- | --- |
| 1 | Team 9 (Invitae Moon) | 9 | 13 | 13 | 13 | 13 | 13 | 92.9 ± 7.3 | 0.64 ± 0.12 | 0.88 | 1 ± 0 | 1 | 2 |
| 1 | Team 12 (Lichtarge) | 9 | 12 | 12 | 12 | 12 | 12 | 85.7 ± 9.3 | 0.65 ± 0.11 | 0.6 | 1.21 ± 0.43 | 2 | 1 |
| 1 | Team 11 (enGenome) | 7 | 10 | 11 | 11 | 11 | 11 | 75 ± 10.5 | 0.49 ± 0.09 | 0.16 | 1.64 ± 1.69 | 3 | 4 |
| 3 | Team 14 (TCS) | 5 | 10 | 11 | 11 | 11 | 11 | 75 ± 10.6 | 0.34 ± 0.1 | 0.8 | 1.07 ± 1 | 3 | 11 |
| 2 | Team 14 (TCS) | 4 | 10 | 11 | 11 | 11 | 11 | 75 ± 11.1 | 0.34 ± 0.1 | 0.8 | 1.07 ± 1 | 3 | 11 |
| 1 | Team 5 (Exomiser) | 6 | 9 | 11 | 12 | 12 | 12 | 73.2 ± 8.8 | 0.47 ± 0.1 | 0.87 | 1.14 ± 1.29 | 6 | 6 |
| 3 | Team 12 (Lichtarge) | 8 | 10 | 10 | 10 | 11 | 11 | 72.1 ± 12.7 | 0.32 ± 0.1 | 0.95 | 1.71 ± 2.05 | 7 | 19 |
| 2 | Team 12 (Lichtarge) | 5 | 10 | 10 | 10 | 11 | 11 | 72.1 ± 11.9 | 0.26 ± 0.09 | 0.95 | 1.79 ± 1.85 | 7 | 25 |
| 3 | Team 11 (enGenome) | 4 | 9 | 11 | 11 | 11 | 12 | 71.8 ± 10.8 | 0.35 ± 0.09 | 0.12 | 1.43 ± 1.65 | 9 | 10 |
| 4 | Team 11 (enGenome) | 7 | 10 | 10 | 10 | 10 | 10 | 71.4 ± 10.8 | 0.48 ± 0.1 | 0.19 | 0.79 ± 0.89 | 10 | 5 |
| 2 | Team 5 (Exomiser) | 6 | 10 | 10 | 10 | 10 | 10 | 71.4 ± 12.3 | 0.53 ± 0.1 | 0.61 | 1.71 ± 1.82 | 10 | 3 |
| 2 | Team 11 (enGenome) | 7 | 9 | 10 | 10 | 10 | 10 | 67.9 ± 12 | 0.43 ± 0.1 | 0.1 | 1 ± 1.47 | 12 | 8 |
| 1 | Team 14 (TCS) | 6 | 9 | 10 | 10 | 10 | 10 | 67.9 ± 12.3 | 0.45 ± 0.11 | 0.9 | 0.57 ± 0.85 | 12 | 7 |
| 3 | Team 5 (Exomiser) | 4 | 9 | 10 | 10 | 10 | 10 | 67.9 ± 12 | 0.34 ± 0.09 | 0.88 | 1.07 ± 1.64 | 12 | 11 |
| 2 | Team 10 | 2 | 9 | 9 | 10 | 11 | 11 | 66.8 ± 12 | 0.26 ± 0.08 | 0.98 | 1.71 ± 0.91 | 15 | 25 |
| 1 | Team 10 | 1 | 6 | 8 | 11 | 11 | 11 | 55.4 ± 10.9 | 0.14 ± 0.06 | 0.99 | 5.07 ± 5.15 | 16 | 39 |
| 2 | Team 1 | 4 | 7 | 7 | 8 | 10 | 10 | 53.2 ± 13.1 | 0.36 ± 0.09 | 0.07 | 1.36 ± 1.22 | 17 | 9 |
| 2 | Team 4 (DITTO) | 3 | 7 | 7 | 8 | 10 | 10 | 53.2 ± 13.4 | 0.32 ± 0.08 | 0.86 | 1.71 ± 2.02 | 17 | 19 |
| 4 | Team 4 (DITTO) | 3 | 7 | 7 | 8 | 10 | 10 | 53.2 ± 12.8 | 0.32 ± 0.08 | 0.86 | 1.64 ± 2.02 | 17 | 19 |
| 1 | Team 4 (DITTO) | 3 | 7 | 7 | 8 | 10 | 10 | 53.2 ± 11.7 | 0.33 ± 0.08 | 0.86 | 1.57 ± 1.74 | 17 | 17 |
| 5 | Team 4 (DITTO) | 3 | 7 | 7 | 8 | 10 | 10 | 53.2 ± 11.7 | 0.33 ± 0.09 | 0.86 | 1.57 ± 1.74 | 17 | 17 |
| 3 | Team 4 (DITTO) | 3 | 7 | 7 | 8 | 10 | 10 | 53.2 ± 12.1 | 0.34 ± 0.09 | 0.86 | 1.5 ± 1.74 | 17 | 11 |
| 6 | Team 4 (DITTO) | 3 | 7 | 7 | 8 | 10 | 10 | 53.2 ± 12.4 | 0.34 ± 0.09 | 0.86 | 1.5 ± 1.74 | 17 | 11 |
| 4 | Team 5 (Exomiser) | 2 | 6 | 8 | 9 | 10 | 10 | 52.5 ± 11.7 | 0.24 ± 0.1 | 0.96 | 1.43 ± 1.45 | 24 | 30 |
| 1 | Team 1 | 4 | 7 | 7 | 7 | 7 | 9 | 50.7 ± 12.6 | 0.34 ± 0.1 | 0.76 | 1.07 ± 0.92 | 25 | 11 |
| 4 | Team 1 | 4 | 7 | 7 | 7 | 7 | 8 | 50.4 ± 15.1 | 0.27 ± 0.11 | 0.99 | 0.57 ± 0.51 | 26 | 22 |
| 1 | Team 2 (AIBI) | 5 | 6 | 7 | 8 | 10 | 10 | 49.6 ± 12.8 | 0.22 ± 0.12 | 0.96 | 0.29 ± 0.61 | 27 | 31 |
| 2 | Team 2 (AIBI) | 5 | 6 | 7 | 8 | 10 | 10 | 49.6 ± 11.6 | 0.22 ± 0.11 | 0.96 | 0.29 ± 0.61 | 27 | 31 |
| 5 | Team 2 (AIBI) | 4 | 6 | 7 | 8 | 10 | 10 | 49.6 ± 12.3 | 0.25 ± 0.13 | 1 | 0.14 ± 0.36 | 27 | 28 |
| 4 | Team 2 (AIBI) | 5 | 6 | 7 | 8 | 9 | 10 | 49.3 ± 11.9 | 0.25 ± 0.12 | 0.88 | 0.14 ± 0.36 | 30 | 28 |
| 3 | Team 1 | 3 | 5 | 7 | 9 | 10 | 10 | 47.1 ± 11.1 | 0.27 ± 0.11 | 1 | 0.57 ± 0.51 | 31 | 22 |
| 5 | Team 1 | 3 | 5 | 7 | 9 | 10 | 10 | 47.1 ± 10.4 | 0.27 ± 0.11 | 1 | 0.57 ± 0.51 | 31 | 22 |
| 1 | Team 13 | 4 | 6 | 7 | 7 | 7 | 7 | 46.4 ± 13.3 | 0.17 ± 0.09 | 0.59 | 1.5 ± 2.98 | 33 | 35 |
| 6 | Team 2 (AIBI) | 5 | 5 | 7 | 8 | 10 | 10 | 46.1 ± 11.4 | 0.22 ± 0.1 | 1 | 0.29 ± 0.83 | 34 | 31 |
| 2 | Team 8 (BORG) | 3 | 5 | 6 | 8 | 12 | 13 | 46.1 ± 10 | 0.26 ± 0.09 | 0.36 | 0.64 ± 1.6 | 34 | 25 |
| 3 | Team 2 (AIBI) | 5 | 5 | 7 | 8 | 9 | 9 | 45.4 ± 11.8 | 0.13 ± 0.12 | 0.98 | 0.07 ± 0.27 | 36 | 41 |
| 2 | Team 15 | 6 | 6 | 6 | 6 | 6 | 6 | 42.9 ± 13.6 | 0.08 ± 0.03 | 0.85 | 9.5 ± 3.88 | 37 | 49 |
| 1 | Team 15 | 6 | 6 | 6 | 6 | 6 | 6 | 42.9 ± 13.8 | 0.14 ± 0.09 | 0.83 | 5.14 ± 4.94 | 37 | 39 |
| 2 | Team 13 | 3 | 5 | 6 | 7 | 7 | 7 | 41.1 ± 12.2 | 0.18 ± 0.08 | 0.73 | 0.57 ± 1.16 | 39 | 34 |
| 4 | Team 8 (BORG) | 0 | 3 | 5 | 7 | 10 | 13 | 35.4 ± 9 | 0.15 ± 0.06 | 0.46 | 0.86 ± 2.07 | 40 | 37 |
| 3 | Team 8 (BORG) | 1 | 3 | 5 | 6 | 10 | 11 | 33.6 ± 9.7 | 0.17 ± 0.1 | 0.55 | 0.64 ± 1.39 | 41 | 35 |
| 1 | Team 16 | 1 | 4 | 5 | 5 | 5 | 5 | 32.1 ± 11.1 | 0.12 ± 0.08 | 0.51 | 0.21 ± 0.43 | 42 | 42 |
| 1 | Team 8 (BORG) | 2 | 3 | 3 | 5 | 8 | 11 | 28.2 ± 9.9 | 0.15 ± 0.08 | 0.38 | 2.86 ± 3.18 | 43 | 37 |
| 5 | Team 5 (Exomiser) | 0 | 3 | 4 | 4 | 5 | 7 | 26.4 ± 10.6 | 0.1 ± 0.06 | 0.98 | 2 ± 2.48 | 44 | 48 |
| 1 | Team 7 (Uniss) | 1 | 2 | 4 | 6 | 6 | 7 | 25.4 ± 9.6 | 0.11 ± 0.07 | 1 | 1.71 ± 1.73 | 45 | 47 |
| 3 | Team 16 | 1 | 2 | 4 | 5 | 6 | 6 | 23.9 ± 9.4 | 0.12 ± 0.09 | 0.51 | 0.21 ± 0.43 | 46 | 42 |
| 2 | Team 16 | 1 | 2 | 4 | 4 | 6 | 7 | 23.2 ± 9.9 | 0.12 ± 0.1 | 0.51 | 0.21 ± 0.43 | 47 | 42 |
| 2 | Team 3 (Bologna Biocomputing Group) | 1 | 2 | 3 | 5 | 6 | 6 | 22.1 ± 8.7 | 0.06 ± 0.04 | 1 | 5.64 ± 8.04 | 48 | 50 |
| 4 | Team 16 | 1 | 2 | 2 | 2 | 2 | 2 | 14.3 ± 10 | 0.12 ± 0.08 | 0.51 | 0.21 ± 0.43 | 49 | 42 |
| 1 | Team 3 (Bologna Biocomputing Group) | 1 | 1 | 1 | 1 | 1 | 1 | 7.1 ± 6.5 | 0.12 ± 0.08 | 1 | 0.21 ± 0.43 | 50 | 42 |
| 1 | Team 6 | 0 | 0 | 0 | 0 | 0 | 0 | 0 ± 0 | NA | NA | NA | 51 | 51 |
| 2 | Team 6 | 0 | 0 | 0 | 0 | 0 | 0 | 0 ± 0 | NA | NA | NA | 52 | 52 |

For each team and model, the number of detected causal variants at rank 1, 1-5, 1-10, 1-20, 1-50, and 1-100 is displayed out of a maximum of 14. The mean rank points (maximum 100) and F-max value (maximum 1) assessment metrics are displayed with the bootstrapped SE (see **Methods**). The F-max producing EPCR values and the mean number of predictions per proband at or above this threshold are displayed with the SD. Model performance is ranked separately for each of the two metrics. SE, standard error; SD, standard deviation; F-max, maximum F-measure; EPCR, estimated probability of causal relationship.

#### Team 9 (Invitae Moon)

The Invitae Moon team submitted one model and predicted the causal variant(s) in 13 of 14 solved families within the top five ranked variants, nine at rank position one. At the F-max producing EPCR threshold, a mean of one variant was prioritized per proband. The model’s performance ranked first by the mean rank points metric and second by F-max. Only one diagnosis was missed, a *de novo* variant in *BICC1* for P21, presenting with unilateral multicystic kidney dysplasia and severe infantile onset neutropenia.

Moon™ (Invitae, San Francisco, CA) is an automated analysis software package developed to prioritize likely causative variants from genome or exome sequencing data. Variant prioritization is achieved by an algorithm incorporating i) the patient’s clinical and sequencing data, ii) parental sequencing data and affected status, iii) curated gene-phenotype associations, and iii) variant annotations, including gnomAD frequency, variant effect predictions, ClinVar submissions and Invitae classifications (internal data). Gene-phenotype associations are maintained in the “Apollo” database by trained genetic scientists at Invitae, and kept up-to-date by daily scanning of the published medical literature for new gene-phenotype associations, followed by manual review and curation of relevant information; HPO terms, the number of patient observations for each HPO, range of disease onset for reported individuals, and the reported inheritance pattern and pathogenic mechanism for the gene. Variants were submitted only for genes that have already been associated with Mendelian disorders in scientific literature. Moon™ is a commercial product available for paid licensed use and was used in an automated fashion.

#### Team 12 (Lichtarge)

The Lichtarge team at the Baylor College of Medicine submitted three models. Their top performing model by both metrics, model 1, predicted the causal variant(s) in 12 of 14 solved families within the top five ranked variants, of which nine were at rank position one. At the F-max producing EPCR threshold, a mean of 1.21 variants were prioritized per proband. The model’s performance ranked second by the mean rank points metric and first by the F-max metric. The model did not identify the causal variant(s) for two probands (P6 and P19).

The Lichtarge team developed scoring systems to prioritize missense, nonsense, and frameshift variants. The team left silent, splicing, and non-coding variants out of their analysis, such as the causal variant of P19. They used the Evolutionary Action method (23) to predict the functional consequences of the missense variants, and accounted for variant call quality, population allele frequency, variant segregation pattern in the families (*de novo*, X-linked dominant males, and autosomal recessive), the ability of each gene to tolerate mutations (unpublished score based on Evolutionary Action), and known gene associations with the patient’s phenotype. Their top performing model, model 1, prioritized the variants according to the predicted probability for loss of gene function, in contrast to models 2 and 3 that prioritized variants, above a threshold for predicted loss of gene function, according to their association to the provided phenotypes. Merging of the variants prioritized for different inheritance modes was performed manually using the predictor’s judgement to provide a single submission. These tools are in-house, involved automated and manual analysis, and are not publicly available at this time; more information can be obtained by contacting the authors.

#### Team 11 (enGenome)

The enGenome team submitted four models. Their top performing model by both metrics, model 1, predicted the causal variant(s) in 10 of 14 solved families within the top five ranked variants, of which seven were at rank position one, and predicted 11 of 14 overall. At the F-max producing EPCR threshold, a mean of 1.64 variants were prioritized per proband. The model did not identify the causal variant(s) for three probands (P6, P21, and P23) in their submission. However, with model 3, the enGenome team identified 12 causative variants of 14 overall.

The enGenome team applied ensemble and linear machine learning classifiers trained on the challenge training set. The features set used to identify the causative variant(s) relies on ACMG/AMP variant pathogenicity, computed through enGenome proprietary variant interpretation software eVai, (24,25), as well as variant quality, family segregation and phenotypic similarity. ACMG/AMP classification is computed only if the gene is associated with at least one condition in databases such as MedGen (https://www.ncbi.nlm.nih.gov/medgen/), Disease Ontology (https://disease-ontology.org/), and Orphanet (https://www.orpha.net/) and phenotypic similarity metrics are computed only when the gene is known to be associated with at least one phenotype. This explains the diagnoses missed by enGenome in the test set (P6 and P23), as both causative genes (*KCND2* and *GNAI1*) were not associated with conditions in these databases when the models were trained. In one additional case (P21), the causative gene was not associated with phenotypes in these databases at the time of the challenge and was identified only by model 3. enGenome’s eVai platform is a commercial product available for paid licensed use and was used in an automated fashion.

#### Team 14 (TCS)

The TCS team submitted three models. Their top performing models by mean rank points, models 2 and 3, predicted the causal variant(s) in 10 of 14 solved families within the top five ranked variants, with a maximum of five at rank position one, and predicted 11 overall. Collectively, the models did not identify the causal variant(s) for three probands (P6, P16, and P24). Their top performing model by F-max value was model 1, prioritizing a mean of 0.6 variants per proband at the F-max producing EPCR threshold.

The TCS team used a combination of in-house tools, “VPR” for variant prioritization and “PRIORI-T” (26) and “GPrio” for gene prioritization. Briefly, variants were ranked based on minor allele frequency (MAF), evolutionary conservation, *in silico* predictions of deleteriousness, and prior disease associations. PRIORI-T queries a rare disease heterogeneous association network with the HPO terms for each proband and outputs a ranked list of genes. GPrio calculates gene scores by two methods. The first is based on HPO-gene correlations reported in the HPO database (https://hpo.jax.org/app/) (11). The second uses the STRING-DB database (https://string-db.org/) (27) to explore indirect hits through interacting genes with relevant HPO correlations. Based on different combinations of the tools, three prediction models were submitted, described in the **Supplemental Materials**. The TCS tools are in-house, involved manual analysis, and are not publicly available at this time; more information can be obtained by contacting the authors.

#### Team 5 (Exomiser)

The Exomiser team submitted five models. Their top performing model by mean rank points, model 1, predicted the causal variant(s) in nine of 14 solved families within the top five ranked variants, of which six were at rank position one, and predicted 12 overall. The model did not identify the causal variant(s) for two probands (P24 and P27) in their submission. Their top performing model by F-max value was model 2, prioritizing a mean of 1.71 variants per proband at the F-max producing EPCR threshold.

The open-source Exomiser tool (version 13.0.0) (28) was run using the latest databases (2109) at time of analysis (Sep 2021), along with a local frequency file generated from 86 non-training samples where AC>1. A maximum of 100 variants per model were returned for all candidates with an Exomiser score >0.2 based on Exomiser’s ranking with no further manual intervention. Model 1 used the recommended default Exomiser settings where high quality (FILTER=PASS in input VCF), rare, segregating, coding variants were prioritized based on minor allele frequency, predicted pathogenicity and the similarity of the patient phenotypes to reference genotype to phenotype knowledge from human disease and model organism databases along with neighbors from the STRING-DB protein-protein association databases (https://string-db.org/) (27). Model 2 used the same settings except only reference human disease knowledge was used. Model 3 extended the model 2 analysis to all variants in the VCF, rather than just the high-quality ones. Model 4 extended the model 2 analysis to allow incomplete penetrance where the prioritized variants can also be present in unaffected family members. Model 5 extended the model 3 analysis to non-coding variants in the genome sequence using the Genomiser variant of Exomiser (29). The two diagnoses missed by model 1 were due to a sex-limited phenotype in one case and a low predicted pathogenicity by REVEL and MVP (30,31) in the other. In the latter case, this variant has now been deposited in ClinVar and would be a top-ranked candidate if rerun due to the ClinVar whitelisting feature of Exomiser. For the three diagnoses ranked outside the top five, two involved disease-gene associations that were in the published literature but not present in OMIM at the time of analysis; these would be highlighted as top-ranking candidates if rerun now (May, 2023). Exomiser is open source and freely available and was used in an automated fashion.

### Reanalysis of solved families

Given the high performance of these models, we reanalyzed the solved families in which models ranked variants higher than the causal variants identified by the RGP team in the answer key, to determine if they may contribute to disease or represent a more likely causal diagnosis; however, no compelling variants were found. To illustrate this, a detailed review of the variants prioritized by one of the top performing teams, Team 9 (Invitae Moon) in four solved probands (P2, P6, P7, and P11) is provided in the **Supplemental Material**.

### Review of “difficult to predict” diagnoses

In genomics-driven diagnostics, failure to recognize causal variants and falsely prioritizing non-causal variants are recognized complications (5,32). We therefore reanalyzed families in the answer key for which predictors consistently failed to prioritize the causal variant(s). Several of these are described below.

The most poorly predicted diagnosis was *KCND2* (c.1207C>G, p.Pro403Ala, ENST00000331113) in P6, a patient presenting with infantile-onset bilateral sensorineural hearing impairment, blindness, retinal dystrophy, hypotonia, chorea, profound global developmental delay, intellectual disability, and dystonia. Across all models, the causal variant was never reported at rank position one, was ranked at position 2-5 by just three models, and was only listed by 26 of 52 models (50%) across all variant predictions. This heterozygous ACMG/AMP LP missense variant in *KCND2* explains the patient’s phenotype (33), is predicted to be deleterious by *in silico* prediction (REVEL 0.84 - PP3 Moderate) (8,30), and is absent from large population databases (gnomAD and TOPMed) (3,34). However, only duo sequencing was available for this family, from the proband and unaffected father; therefore, the *de novo* status of the variant remains unconfirmed. This may hinder models in prioritizing the variant. Indeed, calculating the mean rank points metric separately for families with proband-only or duo data versus those with trio or quad data, demonstrates a significant improvement in model performance with trio or quad data (paired Student’s T-Test p-value 0.00086) (**Figure 2C**). *KCND2* is also not yet reported in the OMIM database as Mendelian-disease associated (last accessed April 2023). Thereby, models limiting their assessment to variants within OMIM reported Mendelian-disease associated genes would fail to prioritize this causal variant. This highlights the importance of OMIM and similar databases to the medical genomics community and the need to be able to represent novel gene-disease associations more rapidly. One such option for laboratories reporting novel Mendelian gene-disease relationships is to deposit them in the Gene Curation Coalition (GenCC) Database (https://thegencc.org/) allowing more rapid dissemination of findings to the community as well as the aggregation of many public and private gene-disease databases (35).

The second most poorly predicted diagnosis was *PI4KA* in P5, a patient presenting with global developmental delay, poor coordination, hypotonia, and spasticity, with an MRI-brain demonstrating cerebral hypomyelination and a dysplastic corpus callosum. Across all models, the two causal variants in this recessive gene were found at position 1-5 in nine models and were only listed by 18 of 52 models (35%) across all submitted variants. The first variant is a P nonsense variant (c.1852C>T, p.Arg618Ter, ENST00000255882; ACMG/AMP criteria applied: PVS1, PM2, PP1, PP3, and PP4). The second is a LP missense variant (c.4990G>A, p.Asp1664Asn, ENST00000255882; ACMG/AMP criteria applied: PP1, PP3, PP4, PM1 Supporting, PM2, PM3). One plausible explanation for the low prediction rate is that this is the only proband in the test set with a recessive diagnosis, requiring the models to jointly prioritize a pair of heterozygous variants. Notably, only 11 of 16 teams included proposed biallelic variants in their submissions, though several cases of recessive inheritance were included in the training set. Moreover, as with the *KCND2* family above, this family was only sequenced as a duo (proband and unaffected father), demonstrating paternal inheritance of the nonsense variant, and requiring the assumption that the missense variant is maternally inherited or *de novo*, on the maternal haplotype, to constitute a recessive diagnosis.

The third most poorly predicted diagnosis was a splice acceptor variant in *CLTC* (c.1534del, p.Val512LeufsTer11, ENST00000621829), a gene associated with intellectual disability in OMIM (MIM: 617854). The proband (P19) presented with global developmental delay, hearing impairment, severely reduced visual acuity, constipation, hyperbilirubinemia, pulmonary arterial hypertension, and intracranial hemorrhage. This variant was ranked at position 1-5 by 13 models and was only listed by 18 of 52 models (35%) across all submitted variants. This *de novo* heterozygous LP splice acceptor variant (ACMG/AMP criteria applied: PS2, PM2, PVS1 Moderate) is predicted to cause a frameshift leading to a premature stop codon 11 amino acids downstream (in exon 10 of 31) in a highly loss-of-function constrained gene and is absent from large population databases. Moreover, since the challenge, the *CLTC* variant has been reported as LP in ClinVar by an independent submitter in association with intellectual disability (ClinVar variation ID: 811442). This variant arises at an acceptor splice site in the gene, thereby outside of the protein-coding region, which may explain the lower level of detection.

Finally, the fourth most poorly predicted diagnosis was *TUBB8* in P27 (c.1039A>G, p.Asn347Asp and c.1033C>T, p.Leu345Phe, ENST00000568584), a female proband sequenced as a quad with her affected female sibling and both unaffected parents. In this family, two causal variants in *TUBB8* were identified, inherited in *cis* from the unaffected father. Carriage of the causal variants by the unaffected father is explained by sex-limited expression of the oocyte maturation defect disease phenotype in females (MIM: 616780). To not exclude these variants as causal, the model would need to take sex-limited expression into consideration or allow for incomplete penetrance. Notably, amongst the 11 models ranking at least one of these causal variants at position one was Exomiser model 4, a model specifically allowing for incomplete penetrance, and all six models from DITTO, that did not take familial segregation into account.

### Patterns in model performance

Following assessment of model performance, the prediction assessor was unblinded to the identity and methods of the participating teams. The wide variability in methodology, spanning stepwise filtering approaches to machine learning and artificial intelligence, did not allow for a comprehensive analysis nor use of statistical tests. However, a qualitative review of the methods demonstrated a pattern of decreasing performance when one or more of the following features were not considered by the model: i) variant call quality; e.g., depth, allele balance, and genotype quality (inclusion of sequence artifacts into submissions), ii) variant allele frequency; e.g., rare in large scale population databases such as gnomAD and TOPMed, iii) variant deleteriousness prediction; e.g., use of *in silico* tools and/or training on reported variants in clinical databases such as ClinVar and HGMD, iv) familial segregation within the provided dataset and inheritance mode of the respective gene, and v) relevance of the putative causal variant(s) to the proband’s phenotype. In some cases, all or most features were considered, yet the model did not identify many diagnostic variants, presumably due to the specific methodology used, information sources, and thresholds selected.

### Summary of variant predictions in unsolved probands

Through reanalysis of the 16 unsolved families, directed by the submitted variant predictions from the top 10 teams, two additional families (12.5%) received a genetic diagnosis. The first, by the detection of a *de novo* splice region variant in *TCF4* (c.1228+3G>T, ENST00000398339), prioritized by eight models in total, submitted by Team 9 (Invitae Moon, model 1 at rank 1), Team 5 (Exomiser, model 1-2 at rank 1 and model 3 at rank 2), and Team 11 (enGenome, model 2 and 4 at rank 1, and model 1 and 3 at rank 2). The second, by the detection of compound heterozygous frameshift (c.706del, p.Arg236GlyfsTer8, ENST00000175506) and deep intronic (c.1137+200_1137+205del, ENST00000175506) variants in *ASNS*, submitted as a biallelic prediction by Team 11 (enGenome, model 1, 2, and 4 at rank 1, and model 3 at rank 2) only. Notably, four additional models from Team 9 (Invitae Moon, model 1, rank 7) and Team 2 (AIBI, model 1, 5, and 6 at rank 83-91) prioritized the *ASNS* frameshift variant only. In both probands, the variant(s) impact on the transcript were functionally validated by RNA sequencing and were returned to the families following confirmation in a CLIA certified laboratory (**Supplemental Table 3**).

In a further six unsolved families, variants in putative novel disease genes were prioritized (**Supplemental Table 3**). For four of the six, a submission had already been made by the RGP team to Matchmaker Exchange (*TPPP* in P9, *KCNH8* in P14, *KLHL13* in P15, and *THAP12* in P18). For the remaining two, new submissions were made (*MRPL54* in P25 and *FRY* in P26). To date, Matchmaker Exchange matches warranting further consideration of these candidate genes have not been received, however, functional studies are underway for some candidates through the GREGoR consortium (https://gregorconsortium.org/). Across the remaining unsolved families, no variants identified were deemed of comparably high interest by the RGP team to pursue by functional studies or submission to Matchmaker Exchange.

Overall, there was more limited concordance in the variant predictions submitted between the top performing models in the unsolved families, compared to the solved families (**Figure 3**); and the vast majority of prioritized variants in the unsolved families did not merit further evaluation after review.

**Figure 3.**
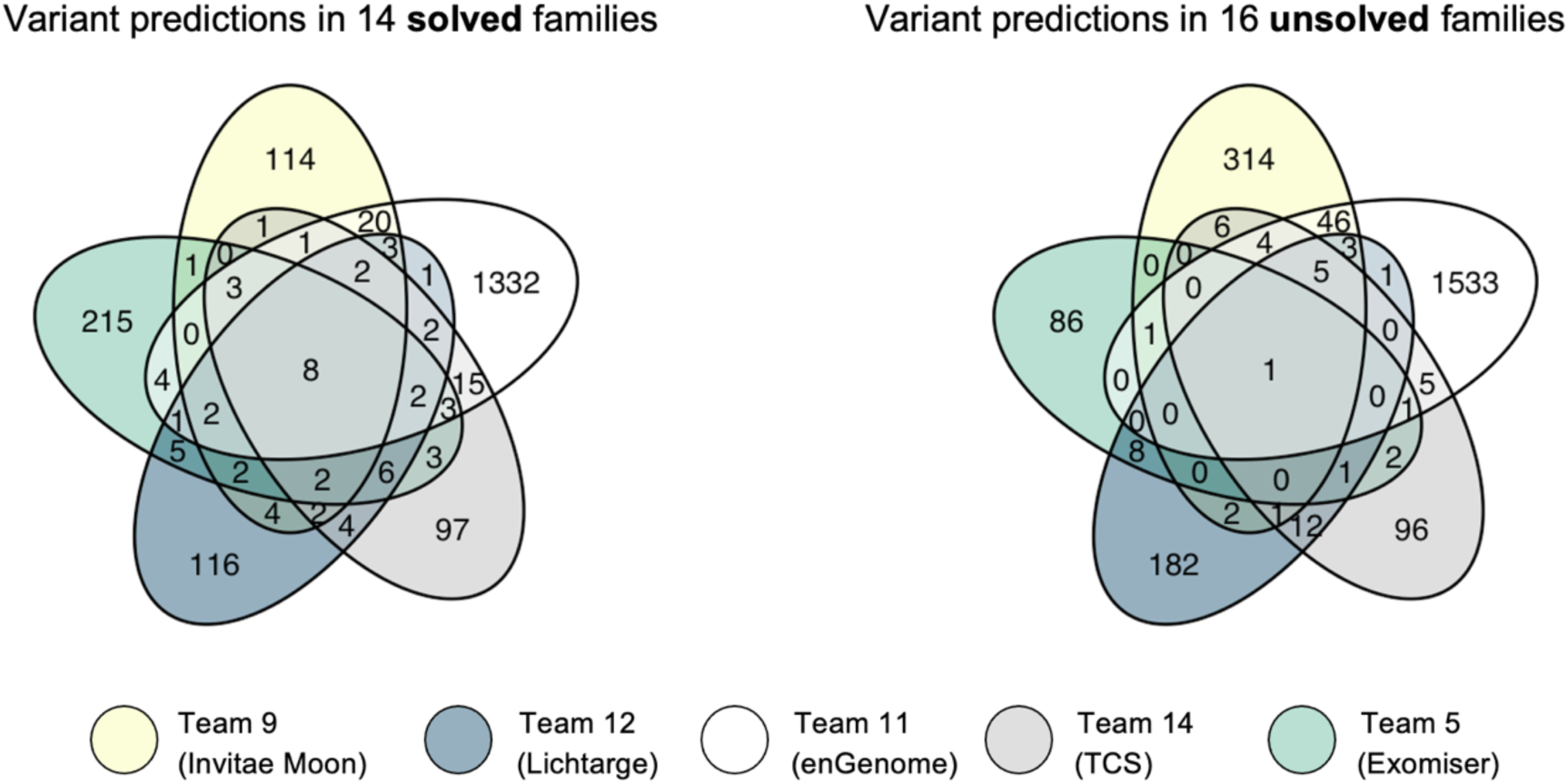
Concordance in the variant predictions submitted by top five performing teams in the solved and unsolved families. Venn diagrams demonstrating the overlap in the variant predictions submitted across all probands in the solved families (left) compared to the unsolved families (right) between top performing teams.

The variants that did not merit further review in the unsolved families mostly fell into one or more of the following categories: i) heterozygous variants in dominant disease genes (according to the reported mode of inheritance in OMIM) inherited from an unaffected biological parent, and where incomplete penetrance is not expected based on current understanding, ii) heterozygous variants in dominant disease genes present in large population databases at an allele frequency higher than consistent with the prevalence of disease, where incomplete penetrance is not expected, and iii) single heterozygous variants in recessive disease genes that are unable to constitute a diagnosis without a second biallelic, compound heterozygous variant. For families with a single recessive variant and at least partial phenotype overlap with the reported phenotype, an SV call set generated by the GATK-SV pipeline was analyzed and the gene was manually reviewed in IGV with the aim to identify an SV in *trans*. This analysis did not, however, result in the detection of any additional variants of interest.

To provide one example, a heterozygous, maternally inherited, missense variant in *GRIK2* was prioritized at rank position one by Team 9 (Invitae Moon) in P15. The variant (c.1066G>A, p.Gly356Arg, ENST00000421544) is predicted to be deleterious by *in silico* predictions (REVEL 0.95 – PP3 Strong) (36) and is absent in large population databases. *GRIK2* is associated with dominant neurodevelopmental delay, impaired language, and ataxia (MIM: 619580) and with recessive intellectual disability (MIM: 611092). The dominant form of disease results from *de novo* gain-of-function variants clustering in a specific domain of ionotropic glutamate receptors, proven to affect channel kinetics and function (37,38). As the *GRIK2* variant prioritized by Team 9 is inherited from the unaffected mother and falls far outside of this functional domain, it is inconsistent with being the cause of dominant disease under the assumption of complete penetrance, whereby every individual who has the variant with show signs and symptoms of the disease. The recessive form of disease results from biallelic loss-of-function variants (39,40). As the proband is lacking a second biallelic variant, the variant can also be deprioritized as a cause of recessive disease.

### Returnable diagnoses identified in two unsolved families

For P1, Team 11 (enGenome) prioritized compound heterozygous putative loss-of-function variants in *ASNS* at rank position 1-3 across four submitted models; a maternally inherited frameshift variant (c.706del, p.Arg236GlyfsTer8, ENST00000175506) and a paternally inherited deep intronic 6 base pair deletion (c.1137+200_1137+205del, ENST00000175506). *ASNS* is a disease gene associated with asparagine synthetase deficiency (MIM: 615574) and is a phenotype match for the proband, who presented with Lennox-Gastaut syndrome, infantile spasms, microcephaly, hypotonia, nystagmus, optic nerve hypoplasia, partial agenesis of the corpus callosum, and delayed myelination. Loss-of-function of *ASNS* is an established disease mechanism in autosomal recessive asparagine synthetase deficiency (41,42). The frameshift variant is rare in large population databases (absent in gnomAD, reported in 1/264,690 alleles in TOPMed) and has recently (Feb, 2022) been reported as P in ClinVar (ClinVar variation ID: 1411238). The variant leads to a premature stop codon in the middle of the gene, in exon six of 13, and is expected to result in a truncated protein. The variant is classified as LP according to ACMG/AMP guidelines (criteria applied: PVS1 and PM2 Supporting). The deep intronic indel between exons 10 and 11 (200bp away from the exon), is absent from large population databases and has a moderate SpliceAI score (0.2) (43) predicting acceptor gain. RNA sequencing analysis performed on blood from the proband demonstrated evidence of complex splice disruption, including intron retention and novel exon creation, leading to a premature stop codon in the middle of the gene (**Figure 4A**). In light of this evidence, the variant was classified as LP according to ACMG/AMP guidelines (criteria applied: PS3, PM3, and PM2 Supporting). *ASNS* was deemed a clinical fit by the family’s local physician. A cerebrospinal fluid (CSF) asparagine level was measured in the proband and was found to be within normal range. Though low CSF asparagine level would further support the diagnosis, normal levels have previously been reported in patients with *ASNS* defects, due to limitations in the sensitivity of the assay (41,44). The family is now pursuing oral asparagine therapy.

**Figure 4.**
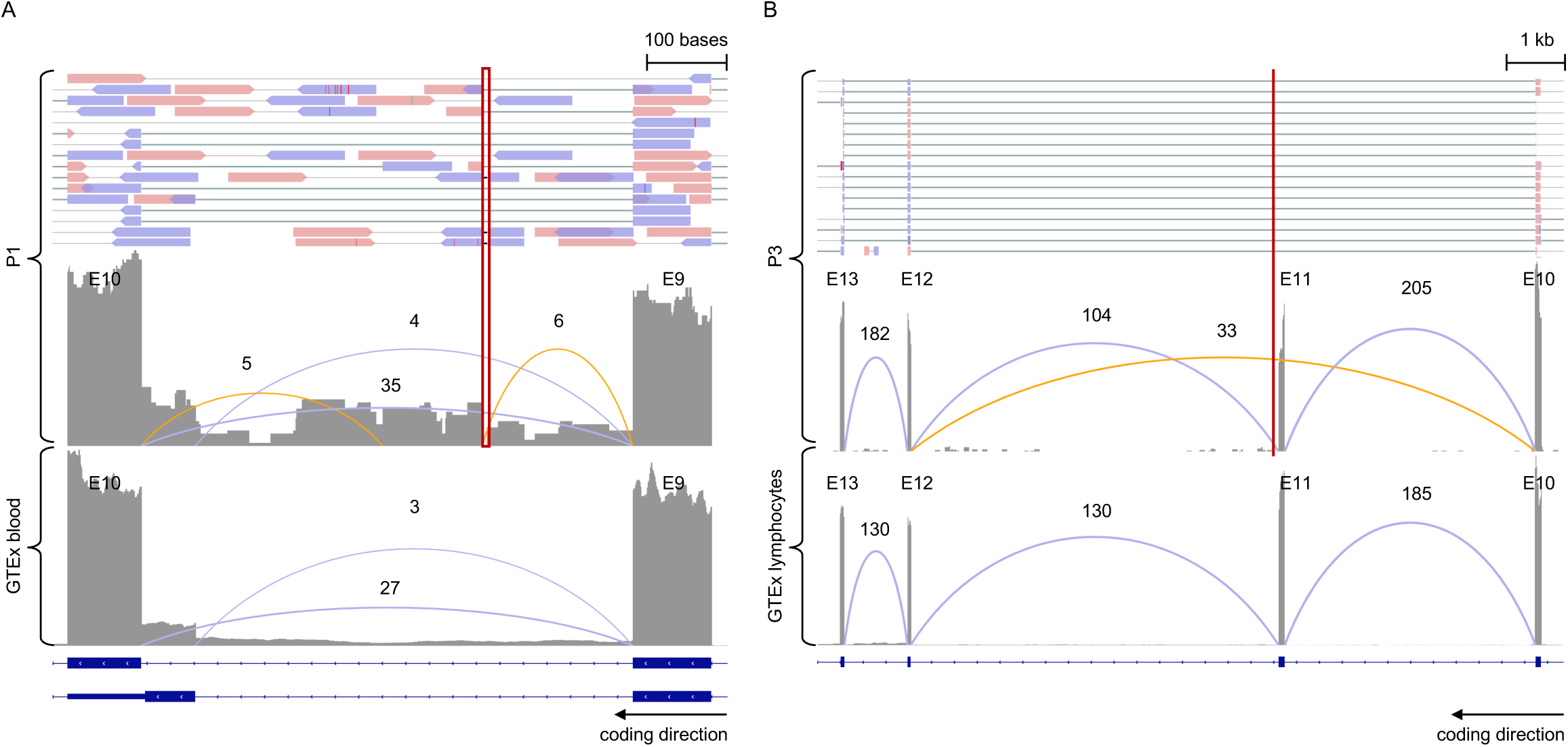
Confirmatory RNA sequencing in P1 and P3. For both **A** and **B**, in the top panel, paired end reads from the RNA sequencing BAM file are displayed for the proband. In the lower panels, the RNA sequencing read pileup tract is displayed with the novel (orange) and known (blue) junctions annotated in the proband and in aggregated data from GTEx controls, respectively. Beneath, the gene transcript isoforms are displayed. **A**, RNA sequencing analysis performed on blood in P1 compared to normalized GTEx blood samples (n=755) (48). The results for *ASNS* (displaying exon 9 and 10) demonstrate evidence of splice disruption due to a deep intronic indel (indicated by the red box in the proband) with cryptic exon creation and intron 9 read-through. **B**, RNA sequencing analysis performed on an EBV-transformed lymphoblastoid cell line (LCL) in P3 compared to normalized GTEx lymphocyte samples (n=174). The results for *TCF4* (displaying exon 10 to 13) demonstrate evidence of splice disruption due to a near-splice variant (indicated by the red line in the proband) with skipping of exon 11 in approximately 20% of reads. E, exon.

In P3, three top performing teams, Team 9 (Invitae Moon), Team 5 (Exomiser), and Team 11 (enGenome), prioritized a *de novo* variant in *TCF4* (c.1228+3G>T, ENST00000398339), a disease gene associated with dominant Pitt-Hopkins syndrome (PHS, MIM 610954). This splice region variant has a moderate SpliceAI score (0.72) predicting donor loss and is absent from large population databases. Moreover, it is a putative loss-of-function variant in a highly loss-of-function constrained gene (pLI score 1, LOEUF 0.22, gnomAD) for which loss-of-function is an established disease mechanism (45). This *TCF4* variant was flagged during analysis in *seqr* by the RGP team. However, at the time, it was considered non-compelling due to the absence of classical PHS features in the proband, such as dysmorphism, including a large beaked nose, wide mouth, fleshy lips, and clubbed fingertips, and abnormal breathing patterns, presenting as hyperventilation episodes. The phenotypic spectrum of *TCF4* has, however, since been expanded to include neurodevelopmental delay in the absence of classical PHS (46). Moreover, upon re-contacting the family for additional clinical information and to request photographs, abnormal breathing patterns and mild dysmorphic features supporting PHS were confirmed. The variant has recently (Aug, 2021) been independently reported in ClinVar as LP (variation ID: 1204043), and has been reported in a study generating patient-specific induced pluripotent stem cells to model PHS (47). RNA sequencing analysis performed on cultured lymphoblasts from the proband demonstrated evidence of splice disruption with exon skipping in the middle of the gene, in exon 11 of 20 (**Figure 4B**). The variant was thereby classified as LP according to ACMG/AMP guidelines (criteria applied: PVS1 and PM2 Supporting).

## DISCUSSION

Genome sequencing allows us to identify the majority of DNA variants in the human genome. However, we are only able to interpret the clinical relevance of a small subset. To aid in bridging this knowledge gap, a broad-spectrum of *in silico* deleteriousness prediction and meta-prediction tools of variant impact have been developed (30,43), and large population databases provide allele frequencies (3,34,49) that enable metrics such as loss-of-function intolerance (50) and missense constraint (51) to be assigned genome-wide. The precise nature in which these tools can most effectively be integrated with phenotype information to pinpoint the genetic diagnosis for an individual patient remains an open question and has spurred the development of numerous computational algorithms integrating machine learning, artificial intelligence, natural language processing, and HPO semantic similarity, among others (9).

The CAGI6 RGP challenge was thereby designed to assess the state of the art in rare disease genome interpretation and to stimulate the development of new methods, providing a forum for highlighting innovation and successes, and discussing bottlenecks. While variants reported by clinical laboratories are often previously observed in patients with the same phenotype or are unreported loss-of-function variants in a gene where loss-of-function is a known mechanism of disease (6,7), we specifically selected solved families with unreported predominantly missense variants, which are often classified as VUS without careful consideration, and included a predominance of families for which no causal variant was yet identified following current field standards. We selected two numeric assessment metrics and found wide variability in model performance. Such wide variability is expected for a challenge encouraging participation from teams experimenting with novel models side-by-side to those with well-established models and infrastructure.

Given that formal variant curation in line with the ACMG/AMP guidelines (6) requires considerable time and is likely to be undertaken only for a handful of highly ranked variants per proband in the clinical setting, for the assessment we developed a simple weighted point allocation metric, the mean rank points metric. This metric did not take into consideration the EPCR values submitted by the teams, and focused only on the rank position of the variants. The metric was weighted to most highly reward models ranking the causal variant(s) at position 1-5, followed by 6-10, 11-20, 21-50, and 51-100, with the number of awarded points falling rapidly for variants ranked in the lower categories. In contrast, our second assessment metric, the F-max value required the models to have a consistent EPCR scoring system across all probands. This metric rewarded models able to stratify causal from non-causal variants at an optimized EPCR threshold, determined by the F-max. This is important as models with a reliable EPCR threshold for the detection of causal variant can support the decision of when an analyst can conclude analysis of a diagnostic genome and deem the result inconclusive, as opposed to arbitrarily manually curating the top 5 or 10 ranked variants, for example.

Overall, we found provision of genome sequencing data from the biological parents and an affected sibling as trios or quads to have a significant positive impact on model performance. This emphasizes the importance of sequencing the biological parents of the proband to ease the analytical burden of variant interpretation by enabling variant phasing and identification of *de novo* variants. Models weighing variant call quality, population allele frequency, and predicted functional consequence in the context of variant segregation in the family, expected inheritance mode of the condition associated with the respective gene, and clinical correlation with the patient’s phenotype, according to current knowledge, were more effective in correctly identifying causal variants at higher rank, as opposed to those where one or more of these features were not taken into consideration. There were minor discrepancies between the performance of the models depending on the metric used for assessment; however, the top performing teams were reasonably consistent. Among the top performing teams was Team 9 (Invitae Moon), identifying all but one diagnosis in the true positive families within the top five rank positions, including two that no other model had ranked as highly. Moreover, three of the top performing teams, Team 11 (enGenome), Team 9 (Invitae Moon), and Team 5 (Exomiser), contributed to the diagnosis of two previously unsolved probands by prioritizing variants that led to return of the result to the families following functional validation through RNA sequencing, formal curation, and clinical confirmation. This included a compound heterozygous diagnosis in *ASNS* in one previously unsolved family, prioritized by Team 11 (enGenome), that indicated a targeted therapy of potential clinical benefit, oral asparagine therapy (52). The large volume of genetic sequencing and phenotype data mined to curate variants and for model development by teams with diagnostic laboratories, may have given an advantage. For example, key considerations enabling the upgrade of a variant from VUS to P/LP; e.g., report of a specific clinical phenotype or identification of a variant in multiple unrelated individuals, may come from internal knowledge and unpublished data.

Looking into the variant predictions in our unsolved families, we found many of the prioritized variants to segregate incorrectly in the family, to have a higher population allele frequency than feasible for the respective Mendelian disease, to be inconsistent with the expected mode of inheritance, to have no clear functional consequence based on currently available *in silico* deleteriousness prediction tools, or to have insufficient overlap with the patient’s phenotype to be considered plausible, raising a number of issues. First, our reanalysis of the unsolved families assumed complete penetrance, unless incomplete penetrance is reported for the gene, and monogenic cause. We therefore deprioritized variants with higher-than-expected allele frequencies that may, arguably, play a role in incompletely penetrant or higher-order oligogenic disease. Second, beyond cases of a clear phenotypic overlap, such as the newly diagnosed *ASNS* proband, that involved a deep intronic indel, we did not consider non-coding variants without *in silico* support of deleteriousness. The strength of some models, for example in recognizing deleterious non-coding variants, may therefore be under-appreciated by the design of this challenge and current knowledge limitations, and would be better positioned to perform highly in a CAGI challenge with a functional read out of variant consequence as the answer key. Functional interpretation of variants remains a major limitation in genome sequencing analysis for which integration of high-throughput functional “omics” data, such as RNA sequencing and quantitative proteomics analyses, and multiplex assays of variant effect (MAVE) (53), including deep mutational scanning, massively parallel reporter assays, and saturation genome editing, will aid in identification of causal variants (54–56). Third, there was an overall scarcity of phenotype overlap between the gene with the prioritized variants and the HPO terms of the respective proband, indicating room for improvement in phenotype matching methodology. For each of these scenarios, it is reasonable to consider that some of the variants identified in the challenge may in the future be reclassified as P/LP if phenotype and mode of inheritance expansions are made and with increased knowledge of incomplete penetrance, to name just a few mechanisms. Notably, some models, including that from Team 9 (Invitae Moon), that limited their analysis of unreported variants to coding regions of known disease genes, and Team 11 (enGenome), that focused only on clinical genes, with established gene-condition and gene-phenotype associations, selected approaches that limit noise at the cost of hindering discovery. It was not a limiting factor for the performance of these teams in the RGP challenge because we did not include gene discoveries in the answer key given it is more difficult to assert an answer is correct in the case of a gene discovery. Finally, many models had poor discriminatory power to prioritize a small number of variants, though this is also due to study design as this was not defined as a goal for submitting candidates. The long lists of candidates underscores the high rate of detection of unreported, unrecognized, findings in genome sequencing data and cautions the overinterpretation of rare variants, and direct usability of these approaches without careful human review. In this regard, though predictive models are approaching useful clinical decision support tools, they require detailed review and conservative assessment of variants against established criteria for use in diagnostics.

The CAGI6 RGP challenge has several limitations. Unlike other CAGI prediction challenges where teams are often tasked to predict functional consequences for variants where the resultant enzyme activity had been quantitatively measured, there was no definitive answer key for this challenge. The answer key used in assessment reflects to the best of our team’s abilities to identify the causal variant(s) in the family following current clinical field standards. We proactively selected families where the causal variant was not reported as disease-causing in ClinVar or HGMD at the time of challenge design, in order to task the models to identify novel causal variants, and delayed submission of the variant to ClinVar for the duration of the challenge. This skewed the spectrum of selected families toward novel heterozygous *de novo* variants and resulted in the inclusion of only one compound heterozygous recessive diagnosis. In future iterations of the challenge, we recommend including a wider array of inheritance modes for diagnoses so that we may give recognition to models that are able to weight other inheritance modes. The challenge was also limited to SNVs and small indel variants, and did not include other classes of variant; e.g., SVs, short tandem repeats, mitochondrial DNA variants, or epigenetic alterations. Each assessment methodology also had limitations and neither precisely models the clinical challenge of identifying variants to be reported, where we want to understand both sensitivity (for discovery) and specificity (for clinical reporting), which are different goals. Predictors were also not informed of the specific scoring used for the rank-based metric nor about the application of the F-max metric and therefore how the choice of assessment method might impact them. However, for the scale of the data, and with subsequent analysis, our selected assessment metrics effectively identified the strength and weakness of different prediction models. As reflected in the methods of some teams, for the CAGI6 RGP challenge, we did not stipulate that entries should not undergo manual curation prior to submission. Therefore, we cannot mitigate the risk of model performance reflecting, to some extent, the result of human review. In future, we intend to offer two challenges, one version with a similar study design to CAGI6 and a second version where we require teams to submit the automated output of the model without human review/reprioritization. We could also ask teams to provide estimates of run time and cost to gain an appreciation for the computational power required and burden of the model. Early participation in RGP was predominantly by families of European descent and that is reflected in the RGP CAGI6 challenge. We hope to have a more US-representative cohort in future challenges and have been working on approaches to diversify participation (20).

Overall, CAGI challenges provide essential information about methods in the field, evaluating both commercial and non-commercial tool performance on unpublished datasets through independent assessment. The CAGI6 RGP challenge has seen among the highest participation of teams to date, in particular increased uptake from industry, even with the higher bar to participate by requiring predictors sign a data use agreement. The challenges are, however, only as good as the amount of participation from research and industry teams, as well as clinical diagnostic laboratories, and involvement is greatly encouraged and appreciated.

## CONCLUSIONS

Computational models for genome analysis were found to be highly variable in terms of methodology and effectiveness. For the diagnosis of patients with rare disease, top performing models significantly aid genome interpretation, and were able to provide new insights into the genetic basis of rare disease by novel variant detection, leading to the return of new diagnostic variants, and to prioritize novel disease genes for further consideration. Overall, we find that computational models can act as clinical decision support tools, requiring detailed review and conservative assessment of prioritized variants against established criteria for use in diagnostics.

## LIST OF ABBREVIATIONS

ACMG/AMP: American College of Medical Genetics and Genomics and the Association for Molecular Pathology
AD: autosomal dominant
AFR: African/African American
AMR: Admixed American
AR: autosomal recessive
ASJ: Ashkenazi Jewish
CAGI: Critical Assessment of Genome Interpretation
CSF: cerebrospinal fluid
DM: disease mutation
EPCR: estimated probability of causal relationship
F-max: maximum F-measure
HPO: Human Phenotype Ontology
IGV: Integrative Genome Viewer
indel: small insertion/deletion
LP: likely pathogenic
NFE: Non-Finnish European
P: pathogenic
PHS: Pitt-Hopkins syndrome
RGP: Rare Genomes Project
SAS: South Asian
SE: standard error
SNV: single nucleotide variant
SV: structural variant
VCF: variant call file
VEP: Variant Effect Predictor
VUS: variant of uncertain significance
XLR: X-linked recessive

## DECLARATIONS

### Ethics approval and consent to participate

The Rare Genomes Project study is approved by the Mass General Brigham Institutional Review Board (IRB) protocol 2016P001422.

### Consent for publication

Written informed consent for the publication of clinical details was obtained from the participants or legal guardians.

### Availability of data and materials

Sequence CRAM files and metadata for the Rare Genomes Project is available through the Broad Institute Data Use Oversight System (DUOS) at duos.broadinstitute.org under dataset IDs DUOS-000008 and DUOS-000143. Contact the authors for the sample ID code used in the CAGI challenge.

### Authors’ contributions

Authors S.L.S., M.O.L., C.B., S.E.B., P.R., H.L.R., and A.O’D-L. designed the challenge/study. Authors S.E.B., P.R., C.B., Y.P., A.K., and S.M.F. organized and provided technical support for the CAGI-RGP challenge. Authors M.A., A.A., G.B., R.B., S.B., P.C., M.G.C., R.C., P.J.C., F.D.P., M.F., M.G., R.H., J.O.B.J., T.J., P.K., C.K., O.L., I.L., Y.L., P.M., T.K.K.M., P.L.M., M.M., G.N., K.N., V.P., Y.P., M.S.P., A.R., E.R., C.S., P.S., Y.S., N.S., D.S., D.S., R.S., Y.S., U.S., W.T., N.T., S.G.V., X.W., Y.W., A.W., E.A.W., R.Y., Y.Y., D.Z., and S.Z. participated as predictors in the CAGI-RGP challenge and provided methods; these authors are sorted alphabetically. Authors S.L.S., M.O.L., G.L., G.R.V., S.D., V.S.G., E.G., E.O.H., B.M., I.O.O., L.P., J.S., M.S-B., B.W., M.W., C.A.T., and P.R. contributed to data analysis and data visualization. Authors S.L.S. and A.O’D-L. wrote the original draft. All authors reviewed and edited the draft and approved the final version.

## Acknowledgements

We thank the many families who participate in the Rare Genomes Project to help improve genetic diagnosis along with the Broad Institute Center for Mendelian Genomics team.

## REFERENCES

1. Splinter K, Adams DR, Bacino CA, Bellen HJ, Bernstein JA, Cheatle-Jarvela AM, et al. Effect of Genetic Diagnosis on Patients with Previously Undiagnosed Disease. N Engl J Med. 2018 Nov 29;379(22):2131–9.

2. 100,000 Genomes Project Pilot Investigators, Smedley D, Smith KR, Martin A, Thomas EA, McDonagh EM, et al. 100,000 Genomes Pilot on Rare-Disease Diagnosis in Health Care - Preliminary Report. N Engl J Med. 2021 Nov 11;385(20):1868–80.

3. Karczewski KJ, Francioli LC, Tiao G, Cummings BB, Alföldi J, Wang Q, et al. The mutational constraint spectrum quantified from variation in 141,456 humans. Nature. 2020 May;581(7809):434–43.

4. Rehm HL. Evolving health care through personal genomics. Nat Rev Genet. 2017 Apr;18(4):259–67.

5. Clark MM, Stark Z, Farnaes L, Tan TY, White SM, Dimmock D, et al. Meta-analysis of the diagnostic and clinical utility of genome and exome sequencing and chromosomal microarray in children with suspected genetic diseases. NPJ Genom Med. 2018 Jul 9;3:16.

6. Richards S, Aziz N, Bale S, Bick D, Das S, Gastier-Foster J, et al. Standards and guidelines for the interpretation of sequence variants: a joint consensus recommendation of the American College of Medical Genetics and Genomics and the Association for Molecular Pathology. Genet Med. 2015 May;17(5):405–24.

7. Abou Tayoun AN, Pesaran T, DiStefano MT, Oza A, Rehm HL, Biesecker LG, et al. Recommendations for interpreting the loss of function PVS1 ACMG/AMP variant criterion. Hum Mutat. 2018 Nov;39(11):1517–24.

8. Pejaver V, Byrne AB, Feng B-J, Pagel KA, Mooney SD, Karchin R, et al. Calibration of computational tools for missense variant pathogenicity classification and ClinGen recommendations for PP3/BP4 criteria. Am J Hum Genet. 2022 Dec 1;109(12):2163– 77.

9. Jacobsen JOB, Kelly C, Cipriani V, Research Consortium GE, Mungall CJ, Reese J, et al. Phenotype-driven approaches to enhance variant prioritization and diagnosis of rare disease. Hum Mutat. 2022 Aug;43(8):1071–81.

10. The Critical Assessment of Genome Interpretation Consortium. CAGI, the Critical Assessment of Genome Interpretation, establishes progress and prospects for computational genetic variant interpretation methods [Internet]. arXiv [q-bio.GN]. 2022. Available from: http://arxiv.org/abs/2205.05897

11. Köhler S, Gargano M, Matentzoglu N, Carmody LC, Lewis-Smith D, Vasilevsky NA, et al. The Human Phenotype Ontology in 2021. Nucleic Acids Res. 2021 Jan 8;49(D1):D1207–17.

12. McKenna A, Hanna M, Banks E, Sivachenko A, Cibulskis K, Kernytsky A, et al. The Genome Analysis Toolkit: a MapReduce framework for analyzing next-generation DNA sequencing data. Genome Res. 2010 Sep;20(9):1297–303.

13. Pais LS, Snow H, Weisburd B, Zhang S, Baxter SM, DiTroia S, et al. seqr: A web-based analysis and collaboration tool for rare disease genomics. Hum Mutat. 2022 Jun;43(6):698–707.

14. Landrum MJ, Lee JM, Benson M, Brown G, Chao C, Chitipiralla S, et al. ClinVar: public archive of interpretations of clinically relevant variants. Nucleic Acids Res. 2016 Jan 4;44(D1):D862–8.

15. Stenson PD, Mort M, Ball EV, Chapman M, Evans K, Azevedo L, et al. The Human Gene Mutation Database (HGMD®): optimizing its use in a clinical diagnostic or research setting. Hum Genet. 2020 Oct;139(10):1197–207.

16. Radivojac P, Clark WT, Oron TR, Schnoes AM, Wittkop T, Sokolov A, et al. A large-scale evaluation of computational protein function prediction. Nat Methods. 2013 Mar;10(3):221–7.

17. Efron B, Tibshirani R. Bootstrap Methods for Standard Errors, Confidence Intervals, and Other Measures of Statistical Accuracy. SSO Schweiz Monatsschr Zahnheilkd. 1986 Feb;1(1):54–75.

18. Collins RL, Brand H, Karczewski KJ, Zhao X, Alföldi J, Francioli LC, et al. A structural variation reference for medical and population genetics. Nature. 2020 May;581(7809):444–51.

19. Robinson JT, Thorvaldsdóttir H, Winckler W, Guttman M, Lander ES, Getz G, et al. Integrative genomics viewer. Nat Biotechnol. 2011 Jan;29(1):24–6.

20. Serrano JG, O’Leary M, VanNoy G, Holm IA, Fraiman YS, Rehm HL, et al. Advancing Understanding of Inequities in Rare Disease Genomics. medRxiv [Internet]. 2023 Mar 29; Available from: http://dx.doi.org/10.1101/2023.03.28.23286936

21. Miller DT, Lee K, Gordon AS, Amendola LM, Adelman K, Bale SJ, et al. Recommendations for reporting of secondary findings in clinical exome and genome sequencing, 2021 update: a policy statement of the American College of Medical Genetics and Genomics (ACMG). Genet Med. 2021 Aug;23(8):1391–8.

22. Dyke SOM, Linden M, Lappalainen I, De Argila JR, Carey K, Lloyd D, et al. Registered access: authorizing data access. Eur J Hum Genet. 2018 Dec;26(12):1721–31.

23. Katsonis P, Lichtarge O. A formal perturbation equation between genotype and phenotype determines the Evolutionary Action of protein-coding variations on fitness. Genome Res. 2014 Dec;24(12):2050–8.

24. Nicora G, Limongelli I, Gambelli P, Memmi M, Malovini A, Mazzanti A, et al. CardioVAI: An automatic implementation of ACMG-AMP variant interpretation guidelines in the diagnosis of cardiovascular diseases. Hum Mutat. 2018 Dec;39(12):1835–46.

25. Nicora G, Zucca S, Limongelli I, Bellazzi R, Magni P. A machine learning approach based on ACMG/AMP guidelines for genomic variant classification and prioritization. Sci Rep. 2022 Feb 15;12(1):2517.

26. Rao A, Joseph T, Saipradeep VG, Kotte S, Sivadasan N, Srinivasan R. PRIORI-T: A tool for rare disease gene prioritization using MEDLINE. PLoS One. 2020 Apr 21;15(4):e0231728.

27. Szklarczyk D, Gable AL, Nastou KC, Lyon D, Kirsch R, Pyysalo S, et al. The STRING database in 2021: customizable protein-protein networks, and functional characterization of user-uploaded gene/measurement sets. Nucleic Acids Res. 2021 Jan 8;49(D1):D605–12.

28. Bone WP, Washington NL, Buske OJ, Adams DR, Davis J, Draper D, et al. Computational evaluation of exome sequence data using human and model organism phenotypes improves diagnostic efficiency. Genet Med. 2016 Jun;18(6):608–17.

29. Smedley D, Schubach M, Jacobsen JOB, Köhler S, Zemojtel T, Spielmann M, et al. A Whole-Genome Analysis Framework for Effective Identification of Pathogenic Regulatory Variants in Mendelian Disease. Am J Hum Genet. 2016 Sep 1;99(3):595– 606.

30. Ioannidis NM, Rothstein JH, Pejaver V, Middha S, McDonnell SK, Baheti S, et al. REVEL: An Ensemble Method for Predicting the Pathogenicity of Rare Missense Variants. Am J Hum Genet. 2016 Oct 6;99(4):877–85.

31. Qi H, Zhang H, Zhao Y, Chen C, Long JJ, Chung WK, et al. MVP predicts the pathogenicity of missense variants by deep learning. Nat Commun. 2021 Jan 21;12(1):510.

32. Shendure J, Findlay GM, Snyder MW. Genomic Medicine-Progress, Pitfalls, and Promise. Cell. 2019 Mar 21;177(1):45–57.

33. Zhang Y, Tachtsidis G, Schob C, Koko M, Hedrich UBS, Lerche H, et al. KCND2 variants associated with global developmental delay differentially impair Kv4.2 channel gating. Hum Mol Genet. 2021 Nov 16;30(23):2300–14.

34. Taliun D, Harris DN, Kessler MD, Carlson J, Szpiech ZA, Torres R, et al. Sequencing of 53,831 diverse genomes from the NHLBI TOPMed Program. Nature. 2021 Feb;590(7845):290–9.

35. DiStefano MT, Goehringer S, Babb L, Alkuraya FS, Amberger J, Amin M, et al. The Gene Curation Coalition: A global effort to harmonize gene-disease evidence resources. Genet Med. 2022 Aug;24(8):1732–42.

36. Kircher M, Witten DM, Jain P, O’Roak BJ, Cooper GM, Shendure J. A general framework for estimating the relative pathogenicity of human genetic variants. Nat Genet. 2014 Mar;46(3):310–5.

37. Guzmán YF, Ramsey K, Stolz JR, Craig DW, Huentelman MJ, Narayanan V, et al. A gain-of-function mutation in the GRIK2 gene causes neurodevelopmental deficits. Neurol Genet. 2017 Feb;3(1):e129.

38. Stolz JR, Foote KM, Veenstra-Knol HE, Pfundt R, Ten Broeke SW, de Leeuw N, et al. Clustered mutations in the GRIK2 kainate receptor subunit gene underlie diverse neurodevelopmental disorders. Am J Hum Genet. 2021 Sep 2;108(9):1692–709.

39. Córdoba M, Rodriguez S, González Morón D, Medina N, Kauffman MA. Expanding the spectrum of Grik2 mutations: intellectual disability, behavioural disorder, epilepsy and dystonia. Clin Genet. 2015 Mar;87(3):293–5.

40. Motazacker MM, Rost BR, Hucho T, Garshasbi M, Kahrizi K, Ullmann R, et al. A defect in the ionotropic glutamate receptor 6 gene (GRIK2) is associated with autosomal recessive mental retardation. Am J Hum Genet. 2007 Oct;81(4):792–8.

41. Seidahmed MZ, Salih MA, Abdulbasit OB, Samadi A, Al Hussien K, Miqdad AM, et al. Hyperekplexia, microcephaly and simplified gyral pattern caused by novel ASNS mutations, case report. BMC Neurol. 2016 Jul 15;16:105.

42. Schleinitz D, Seidel A, Stassart R, Klammt J, Hirrlinger PG, Winkler U, et al. Novel Mutations in the Asparagine Synthetase Gene (ASNS) Associated With Microcephaly. Front Genet. 2018 Jul 13;9:245.

43. Jaganathan K, Kyriazopoulou Panagiotopoulou S, McRae JF, Darbandi SF, Knowles D, Li YI, et al. Predicting Splicing from Primary Sequence with Deep Learning. Cell. 2019 Jan 24;176(3):535–548.e24.

44. Ruzzo EK, Capo-Chichi J-M, Ben-Zeev B, Chitayat D, Mao H, Pappas AL, et al. Deficiency of asparagine synthetase causes congenital microcephaly and a progressive form of encephalopathy. Neuron. 2013 Oct 16;80(2):429–41.

45. Giurgea I, Missirian C, Cacciagli P, Whalen S, Fredriksen T, Gaillon T, et al. TCF4 deletions in Pitt-Hopkins Syndrome. Hum Mutat. 2008 Nov;29(11):E242–51.

46. Kalscheuer VM, Feenstra I, Van Ravenswaaij-Arts CMA, Smeets DFCM, Menzel C, Ullmann R, et al. Disruption of the TCF4 gene in a girl with mental retardation but without the classical Pitt-Hopkins syndrome. Am J Med Genet A. 2008 Aug 15;146A(16):2053–9.

47. Sripathy SR, Wang Y, Moses RL, Fatemi A, Batista DA, Maher BJ. Generation of 10 patient-specific induced pluripotent stem cells (iPSCs) to model Pitt-Hopkins Syndrome. Stem Cell Res. 2020 Oct;48:102001.

48. GTEx Consortium. The Genotype-Tissue Expression (GTEx) project. Nat Genet. 2013 Jun;45(6):580–5.

49. 1000 Genomes Project Consortium, Auton A, Brooks LD, Durbin RM, Garrison EP, Kang HM, et al. A global reference for human genetic variation. Nature. 2015 Oct 1;526(7571):68–74.

50. Lek M, Karczewski KJ, Minikel EV, Samocha KE, Banks E, Fennell T, et al. Analysis of protein-coding genetic variation in 60,706 humans. Nature. 2016 Aug 18;536(7616):285–91.

51. Samocha KE, Robinson EB, Sanders SJ, Stevens C, Sabo A, McGrath LM, et al. A framework for the interpretation of de novo mutation in human disease. Nat Genet. 2014 Sep;46(9):944–50.

52. Sprute R, Ardicli D, Oguz KK, Malenica-Mandel A, Daimagüler H-S, Koy A, et al. Clinical outcomes of two patients with a novel pathogenic variant in ASNS: response to asparagine supplementation and review of the literature. Hum Genome Var. 2019 May 22;6:24.

53. Esposito D, Weile J, Shendure J, Starita LM, Papenfuss AT, Roth FP, et al. MaveDB: an open-source platform to distribute and interpret data from multiplexed assays of variant effect. Genome Biol. 2019 Nov 4;20(1):223.

54. Cummings BB, Marshall JL, Tukiainen T, Lek M, Donkervoort S, Foley AR, et al. Improving genetic diagnosis in Mendelian disease with transcriptome sequencing. Sci Transl Med [Internet]. 2017 Apr 19;9(386). Available from: http://dx.doi.org/10.1126/scitranslmed.aal5209

55. Kremer LS, Bader DM, Mertes C, Kopajtich R, Pichler G, Iuso A, et al. Genetic diagnosis of Mendelian disorders via RNA sequencing. Nat Commun. 2017 Jun 12;8:15824.

56. Yépez VA, Gusic M, Kopajtich R, Mertes C, Smith NH, Alston CL, et al. Clinical implementation of RNA sequencing for Mendelian disease diagnostics. Genome Med. 2022 Apr 5;14(1):38.

