## Supplemental Material for "Critical assessment of variant prioritization methods for rare disease diagnosis within the Rare Genomes Project"

### Supplemental Information

### Methodology of participating teams

#### Team 1 Methods

Anonymous, no methods provided.

#### Team 2 (AIBI) Methods

Contact: Yang Shen, Texas A&M University,

We trained machine learning models to predict the probability that an input variant is causal for the paired, input phenotype, using the training data provided. *Variant filtering.* Following CHESS (1), we only retained rare variants with Minor Allele Frequency (MAF) below 0.05 and with VEP-predicted coding consequences (2). *Variant featurization.* Input features for variants were predictions from REVEL (3), gene prioritization scores from Phenolyzer (4), and lineage information (4D from either parent); where missing values were simply replaced by zeros. *Phenotype embedding.* Input features for phenotypes were embeddings using pre-trained BioBERT(5) to embed the descriptive texts of each phenotype node followed by possible graph embedding of the Human Phenotype Ontology. *Model training.* We built a multi-layer perceptron and trained it with five-fold cross validation where the loss function is binary cross entropy. The primary model was an ensemble over the combination of five folds and three phenotype embedding strategies.

**Team 3 (Bologna Biocomputing Group) Methods**

Contact: Giulia Babbi, University of Bologna,

The method of the Bologna Biocomputing Group consists of 4 steps that take into consideration: i) the prediction of the variant effect, ii) the variant allele frequency, iii) the inheritance pattern, iv) the relevance to proband phenotype. Variants are mapped on the genome with the Ensembl Variant Effect Predictor (VEP) (2). We retain only variants affecting the protein-coding regions (i.e., stop gain, frameshift, and missense) and exclude variants with allele frequency > 1%, as derived from gnomAD (https://gnomad.broadinstitute.org/).

We analyze the variants considering the genotypes of the proband's parents/sibling, when provided. To prioritize the most probable causative variants (CVs), we mainly focus on de novo mutations, and homozygous alleles in proband with heterozygous parents.

We collect the genes associated with the clinical phenotypes of each proband from two databases eDGAR (6) and PhenPath (7) that collect and organize data on gene-disease and gene-disease-phenotype relationships, respectively. We then provide the functional characterization of diseases and phenotypes by means of functional enrichments computed with NetGE-PLUS (8).

Model 1 considers only variants on genes directly associated to clinical phenotypes.

For Model 2, we enlarge the set of candidate genes, by including i) genes associated with diseases endowed with the proband phenotypes, and ii) genes that participate to the GO-biological processes associated with the phenotypes.

**Team 4 (DITTO) Methods**

Contact: Manavalan Gajapathy, The University of Alabama at Birmingham,

Our machine learning model (DITTO) was trained using pathogenic and benign non-synonymous single nucleotide variants (nsSNVs) retrieved from ClinVar and HGMD, to predict variant pathogenicity classifications. We utilized VEP (Variant Effect Predictor) to annotate variants with allele frequencies from gnomAD, conservation scores and damage predictions from dbNSFP. To improve the performance of our model, we selected a subset of classifiers from the scikit-learn package and stacked them into a single classifier. This approach allowed us to incorporate multiple methods and improve the learning performance of our model. We also used hiPHIVE (Exomiser) to prioritize genes based on phenotype terms extracted from each sample. The tool outputs a score for each gene based on the match to the phenotype terms provided. We normalized these scores to a (0,1) scale to bring both the Exomiser score and deleterious probability to the same scale. We then calculated the mean value between the normalized Exomiser score and deleterious probability for each variant to prioritize variants for each proband. We did not take familial segregation into account for our predictions.

**Team 5 (Exomiser) Methods.**

Contact: Damian Smedley, Queen Mary University of London,

See main text for Exomiser methods.

### Team 6 Methods

Anonymous, no methods provided.

### Team 7 (Uniss) Methods

Contact: Matteo Floris, University of Sassari,

The algorithm developed by the Uniss Team uses a similarity metric between the HPO terms of the proband's symptoms and the HPO terms of each of the diseases (calculated with the R library 'ontologySimilarity' (9)) associated by Orphanet (Orphadata (10,11)) with the genes with the most deleterious mutations found in the proband. The mutated genes in each proband are then sorted according to the similarity between the HPO terms of the associated diseases and the HPOs of the proband's symptoms.

### Team 8 (BORG) Methods

Contact: Azza Althagafi, King Abdullah University of Science and Technology (KAUST),

*Data preprocessing and annotations.* We performed quality control on the variants with a variant quality threshold of 20. Additionally, we preprocessed the data by normalizing indels, verifying if the reference alleles matched the reference, and splitting multiallelic sites into multiple rows. We also used BCFtools (12) and VCFtools (13) to recover multiallelic variants from multiple rows. To identify and prioritize candidate variants before clinical interpretation, we subjected the final list of variant calls to variant filtering. For this purpose, we developed a custom pipeline based on ANNOVAR (14). We filtered out common variants with a Minor Allele Frequency (MAF) value higher than 1%

in any of the known databases, including the 1000 Genomes frequencies, ExAC (15) for all populations, and gnomAD (16). We then filtered the remaining variants using the family pedigree based on the suspected mode of inheritance. After applying the genotype filters, we annotated the variants using VEP (17) and precalculated CADD score (18). We chose to use CADD for variant pathogenicity prediction in this challenge due to its better performance compared to most other prediction methods, as demonstrated in previous studies (18).

*Family-based Filtering.* To choose the most suitable mode of inheritance for each case, we studied both the training set and the actual testing set for all possibilities while considering the ethnicity. Consequently, in some cases, we prioritized some mode of inheritance filters over others. We use ethnicity to prioritize a family filter based on the recessive mode of inheritance when we suspect likely consanguinity (i.e., we use the amount of consanguinity within an ethnic group as a prior when selecting the mode of inheritance filter to apply). For the family-based filtering, we utilized the recently published method Slivar (19). The method explores practical guidelines for variant (SNP and INDEL) filtering and reports the expected number of candidates for de novo dominant, recessive, and autosomal dominant modes of inheritance. We evaluated different settings and configurations based on the family pedigrees. Using Slivar, for the trios or quads, and duos: we us *segregating_denovo*, *segregating_recessive*, and compound heterozygous *compound-hets* filtering. For the proband only cases: we used *segregating_dominant*, *segregating_recessive* filtering for the variants.

*Causal variants prediction.* After filtering variants, our approach for predicting the causative variant(s) is by combining two main sources of information; the first utilizes the genomic features and pathogenicity prediction using CADD, and the second is based on the phenotype annotations for the affected families combined with the ontology-based machine learning method DL2vec (20). We made four submissions based on the different gene-phenotype representations using DL2vec. We mainly utilize three types of gene annotation features for supervised learning as they perform best in our previous experiments (20): Gene Ontology (GO) (21), Mammalian Phenotype Ontology (MP) (22), and the Human Phenotype Ontology (HPO) (23). Specifically, we obtain the annotations of human genes with functions and cellular locations encoded by the GO, and the phenotypes of their mouse orthologs from the Mouse Genome Informatics (MGI) database and characterized using the MP, and the phenotypes of the human genes using HPO. Furthermore, we obtain phenotype annotations of human diseases with the HPO, in addition to the phenotypes obtained from the training set. To combine the annotations using the different ontologies, we use the integrated PhenomeNET ontology (24). We jointly embed the gene and disease, their ontology-based annotations, and the ontologies used in the annotations in a vector space. We generate embeddings individually using GO, MP, and HP annotations, and their union. We then use a pointwise learning-to-rank model to prioritize gene--disease pairs based on gene--disease associations in the Online Mendelian Inheritance in Men (OMIM) database (25), and the phenotypes in our training set. Our model is based on neural networks; given a pair of embedding vectors *G* and *D* as input, the model independently transforms the embeddings into a lower-dimensional representations using two fully-connected hidden layers, and then computes the inner product followed by a sigmoid function that outputs a value between 0 and 1, and which we use as the prediction score for an association between *G* and *D*. We combine DL2Vec predictions with CADD predictions, and include the top 100 predicted variants with the highest weighted prediction scores using DL2vec phenotype and CADD pathogenicity score for all the submissions.

**Team 9 (Invitae Moon) Methods**

Contact: Keith Nykamp, Invitae,

The automated analysis of CAGI6 test cases was performed using Moon 3.4.0 (Invitae). Annotation sources and versions used were ClinVar (2021-06-09), dbNSFP (4.0), dbscSNV (1.1), Apollo (2021-09-27), gnomAD (3.1) and HPO (2021-02-08). VCF files of both proband and family members (if any), sex and age of onset provided for the proband, the proband's phenotype as HPO terms, and family information were used as input for each analysis. Given Moon's focus on diagnosing known rare genetic disorders, the output only contains variants in genes that have already been associated with Mendelian disorders in scientific literature.

Moon generated a list of potential provisional diagnoses by sequentially annotating filtering and ranking variants. In the first step, low quality variants (as determined based on requirements for AD, DP, QUAL, GQ and allelic imbalance) were excluded. Subsequently, common variants in the population (>2% in gnomAD) were filtered out, except for a subset of more common variants with known pathogenic classification. Variants with either Benign or Likely Benign classifications in ClinVar were also excluded from the analysis. With regard to variant effect, only variants in protein or RNA coding regions, splice site regions and known pathogenic variants in non-coding regions were retained. In family analyses, co-segregation of the variant with the phenotype (i.e., healthy or affected status of included family members), according to autosomal dominant, autosomal recessive, X-linked dominant or X-linked recessive inheritance patterns, was taken into account. Segregation was not only applied as a strict filter criterion, thereby also ensuring that causal mutations following non-Mendelian inheritance (e.g., with incomplete penetrance) would be identified in family analyses.

For each variant, a disorder was annotated based on overlap of the proband's phenotype with known gene-disease associations, and phenotype overlap was scored. Only variants with sufficient phenotype overlap between the proband's phenotype and the annotated disorder were retained. This automated phenotype assessment in Moon is driven by a proprietary gene-disease database called Apollo. Natural language processing of the genetics literature guides selection of relevant gene-disease information, which is added to the Apollo database after manual curation.

Based on the inheritance pattern of the annotated disorder by Moon, additional variable frequency thresholds were applied. In addition, only variants for which the zygosity of the called variant fit the inheritance pattern of the annotated disease were retained in the 'SNV' shortlist. Heterozygous variants in genes annotated with recessive conditions were outputted in a separate 'Carrier' list. In both output lists, variants are ranked based on an analysis-specific p-value that is calculated based on a combination of clinical (e.g., phenotype overlap, age of onset) and genetic annotations (e.g., variant specific properties). This p-value is not an absolute probability of causality, but rather a relative probability which only has relevance within a specific case. However, the Moon p-value was converted into an absolute p-value for submission of the results. A standard deviation, however, could not be provided.

**Team 10 Methods**

Anonymous, no methods provided.

**Team 11 (enGenome) Methods**

Contact: Susanna Zucca, enGenome Srl,

The models submitted by the enGenome team are machine learning classifiers that were selected during training phase through a "Leave-one-Proband-Out" (LOPO) cross-validation. The LOPO cross-validation is carried out on the 35 training probands (with known causatives) according to the following procedure: for each training proband, that proband was considered as the "LOPO test proband", and different machine learning models with different hyper-parameters configurations are trained to predict the causative variants on the remaining 34 training probands. The models are evaluated on the current "LOPO test proband" and the ranking positions of the causative variant are recorded. This procedure is repeated for each training proband. Different machine learning models were evaluated, such as Naïve Bayes, Ensemble methods and Neural Network. The models with better ranking performance were selected, and they were trained on the entire CAGI training set of 35 training probands. For each test proband, the trained models were exploited to prioritize variants according to the machine learning predicted probabilities. Following the challenge, the approach has been further improved to account for refined gene-phenotype and gene-condition relations. A new model trained on a larger training dataset has been integrated in the eVai platform (https://evai.engenome.com/) as a new functionality called "Suggested Diagnosis".

**Team 12 (Lichtarge) Methods**

Contact: Panagiotis Katsonis, Baylor College of Medicine,

*Model 1.* We followed three causal variant segregation patterns in the families that led to two different scoring systems. For de novo causality, the variant should be found in the proband and not found in any of the parents. When parent sequencing data were not available, we asked that the causal variant should not be listed in gnomAD or UK biobank databases. The de novo score was calculated as $sc = QC \cdot SR \cdot FR \cdot EA \cdot LAF \cdot GT \cdot AS$, where *QC* is the filter status (=1 if filter was PASS and =0 otherwise), *SR* is the number of supporting reads, *FR* is the fraction of the supporting reads, *EA* is the variant impact (Evolutionary Action score for missense variants, 100 for nonsense and frame-shift insertion/deletion variants), *LAF* is the negative logarithm of the maximum allele frequency value in gnomAD (26) or UK biobank (27) databases, *GT* is the ability of the gene to tolerate mutations (custom calculation using Evolutionary Action and the gnomAD database), and *AS* is the number of HPO terms associated with the gene according to HPO (28), DisGeNet (29), ClinVar (30), HumSavar (https://www.uniprot.org/docs/humsavar), and VarElect NGS Phenotyper (31). The recessive score was calculated for each gene as $sc = Vm \cdot Vf \cdot GT \cdot AS$, where *Vm* and *Vf* are the largest variant scores, $V = QC \cdot EA \cdot LAF$, inherited by mother and father, respectively. For male patients, we used $Vf = Vm$ for all variants of the X chromosome, in order to account for X-linked dominant patterns. The two scoring systems of *de novo* and recessive analysis had different scales, so they were manually merged for the needs of this challenge according to the predictor's judgement.

*Models 2 and 3.* Unlike model 1 that filtered out genes without known associations to the phenotype of interest and then prioritized variants according to their functional impact, these models filtered out variants with low functional impact and prioritized according to the phenotypic associations, using the VarElect NGS Phenotyper (model 2) and also HPO and ClinVar (model 3). We only considered variants with Evolutionary Action scores above 30 and fraction of supporting reads above 0.05. These variants were prioritized for their gene-phenotype associations.

**Team 13 Methods**

Contact: Daniel Zeiberg, Khoury College of Computer Sciences, Northeastern University, Boston, MA, USA,

Our approach combines variant-pathogenicity prediction with gene-phenotype association inference using a simple probabilistic framework (32) to score variants found in each patient's genome. We then aggregate these combined scores over the different phenotypes observed in each individual to obtain a single score that is used to rank variants in decreasing order of their putative causal roles. Our end-to-end data-driven approach is exploratory, and we focus solely on missense variants due to their putative roles in rare genetic disorders and the challenges involved in asserting their pathogenicity.

*Inferring missense variant pathogenicity.* To score all missense variants in each genome from the test set provided, we used precomputed scores from the dbNSFP database (33) for MutPred (34) (Model 1) and REVEL (3) (Model 2). Variants with missing scores were excluded.

*Inferring gene-phenotype associations using protein-protein interaction networks.* A naive label propagation algorithm was implemented that took as inputs: i) a list of 'seed' genes known to be associated with a given Human Phenotype Ontology (HPO) term (23), and ii) a protein-protein interaction network. Here, we used known associations in HPO to the specific phenotypes of interest in the test set for the former and the STRING protein-protein interaction (PPI) network (35) for the latter. For each gene/HPO term relation in the annotations, MyGene.Info (36) was used to map the Entrez Gene ID to a set of Ensembl protein IDs to ensure compatibility with the STRING PPI network. The label propagation algorithm resulted in a matrix of protein-disease (gene-disease) associations for all proteins in STRING and all HPO terms in the CAGI test set. Each association was assigned a score between zero and one, with one indicating a higher propensity for gene-disease association.

*Variant pre-processing, selection, and ranking.* For each proband, the set of missense variants were filtered by keeping only "PASS" quality variants with allele frequencies, generated from gnomAD (37), less than 0.001. These variants were then cross-referenced against those in the respective parents to include only those variants that were unique to the proband. For each of these variants, the set of gene-disease association scores from the label propagation algorithm were then extracted using the variant's corresponding gene/protein and all HPO terms present in the test set metadata. Each variant's MutPred score was multiplied by the set of protein/HPO association scores, reporting the average and standard deviation of these products. For the secondary submission, this procedure was repeated using REVEL.

**Team 14 (TCS) Methods**

Contact: Aditya Rao, TCS Research,

All variants in the VCF were annotated and scored with the VPR pipeline. Only variants marked "PASS", with GQ >= 30, and VPR scores greater than 0.4 were considered for further processing. Further filtration was performed based on phase data if available and genes with all high scoring variants shared with a parent without disease phenotype discarded. Variant based gene ranks were generated by reverse sorting the variants based on VPR score.

Combined ranked variants in the submission for model 1 were between 6 to 35 (median 16) genes per proband. VPR ranked variants were 66 to 400 (median 95) per sample with corresponding genes ranging from 46 to 366 (median 68) per sample.

*VPR.* The Variant PRioritization (VPR) tool is a rules-based engine that annotates and scores input variants independent of the gene of occurrence based on MAF (minor allele frequency), evolutionary conservation, *in silico* predictions and prior disease associations. The variant score, ranging from 0 to 1, is a weighted sum of individual conservation and functional components. The conservation and functional components in turn are built of block scores from MAF data, conservation scores from different predictors, deleteriousness effect predictors, and custom predictions based on the genomic region of the variant. Each block component score is computed through a voting scheme with each predictor voting for the variant being tolerated (0), probably damaging (0.5) or damaging (1). Individual cutoffs for the vote are set for each predictor and participation is based on prior performance of the predictor on the Clinvar (38) database. If prediction is unavailable for a particular variant, the missing predictions are ignored while computing the block score. For the CAGI RGP challenge, the following predictors were used:

- Conservation: GERP RS, PhyloP30, PhyloP100, PhastCons30, PhastCons100
- MAF: 1000 Genomes, gnomAD (WG and Exomes), alpha, ExAC
- Effect Prediction: CADD_phred, SIFT, SIFT4G, MutPred, MVP, MPC, PrimateAI, MutationTaster, MutationAssessor, FATHMM, REVEL, PROVEAN, MetaSVM, MetaLR, M_CAP

Variant Score = 0.25 * Block Score(Conservation Predictors) + 0.25 * BlockScore(MAF) + 0.5 * Block Score(Effect Predictors

*GPrio.* GPrio is a gene prioritization module that provides gene scores, ranks and indirect gene associations given a set of HPO terms for a proband. The module is built on 2 different methods:

*Method 1.* This method is based on the HPO – Gene correlations downloaded from the HPO (23) database. Based on all the HPO terms for each proband, scores were given as follows:

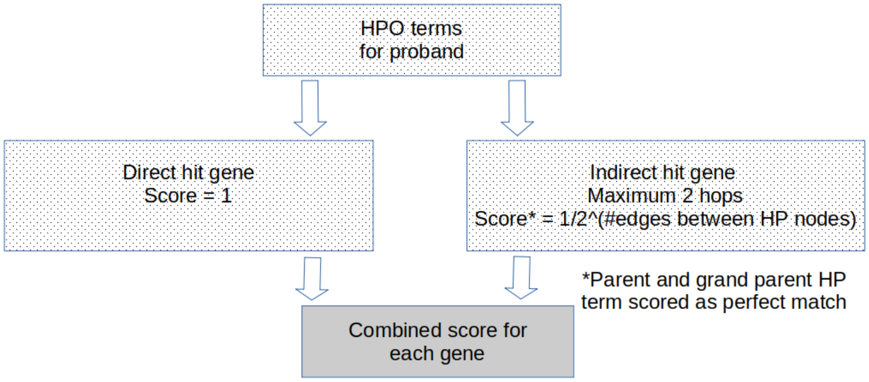

- Direct gene association: 1
- Indirect association with maximum 2 hops:
    - 1 / 2^(number of edges between the HP nodes)
    - In the above case, parent and grandparent HP terms were scored as perfect match

Scores for each gene were summed for all the proband's HPO terms and ranked according to score. For example, if patient has the 2 phenotypes:

- HP:0001324 – Muscle weakness
- HP:0006785 – Limb-girdle muscular dystrophy

Score for the gene TPM2 is computed for the 2 HPO terms as:

- HP:0001324: direct association
- HP:0006785<-HP:0003797 (Limb-girdle muscle atrophy)<-HP:000912 (Abnormal axial skeleton morphology)*
- Hence score is 1 + 1/8 = 1.25

* Grandfather node has direct association

*Method 2.* This method is based on the data from StringDB (39) and all gene – gene interactions with a confidence of above 0.9 are considered. This method is primarily built to explore possible indirect interactions of novel genes without apparent correlation, shortlisted through variant prioritization, with genes that have direct correlations with phenotypes of interest. The score for a gene B is computed as follows:

- For a gene A with a direct correlation with a HPO term, if B is an interacting partner with maximum 2 hops between genes A and B
- Score = Min(Gene-Gene interaction score on path between A and B)

- Combined score for each gene is computed

For example, for a proband with 6 HPO terms, the score for gene GNAI1 is:

- HP:0100704 (['GNAI1', 'GNB1'], 0.999)
- HP:0012448 (['GNAI1', 'GNB1'], 0.999)
- HP:0001290 (['GNAI1', 'GNB1'], 0.999)
- HP:0001263 (['GNAI1', 'GNB1'], 0.999)
- HP:0032807 NOT_FOUND
- HP:0001344 (['GNAI1', 'GNAO1'], 0.968)

Thus 5/6 HPO terms had confident interacting partners and the gene could be a novel association with the disease.

Based on different combinations of these tools, three prediction models were submitted to the CAGI RGP challenge as follows: In model 1, the ranked variants were further filtered by quality and haplotype, where parent data was available, and those variants that overlapped with gene lists generated by either PRIORI-T or GPrio were retained. In addition, high scoring variants from genes that had associations with the proband's phenotypes reported by GPrio's second method or found to have associations through manual curation were also retained. In model 2, the ranked genes and variants from PRIORI-T and VPR, respectively, were combined using an internal combination algorithm to obtain a final ranked list of possible causal genes. This model used no manual intervention. Model 3 was an extension of model 2 with additional manual curation.

**Team 15 Methods**

Anonymous, no methods provided.

**Team 16 Methods**

Anonymous, no methods provided.

**Reanalysis of highly ranked variants by top performing teams in solved families**

In P2, a proband-only family, the Invitae Moon team ranked the causal variant at position two, below proposed biallelic variants in *MYH2*, a ClinVar reported pathogenic frameshift variant (c.1719del, p.Gly574AlafsTer9, ENST00000245503) and a missense variant (c.2390G>T, p.Arg797Met, ENST00000245503) with non-deleterious *in silico* prediction of deleteriousness (REVEL 0.14 – BP4 Moderate) (40). There is a phenotype overlap between *MYH2* and muscle weakness in the proband (MIM 160740). However, variants in *MYH2* do not explain the proband's contractures and pulmonary fibrosis. Moreover, without parental sequencing, these variants cannot be confirmed to be in *trans*. In contrast, the missense variant in *FAM111B* in the answer key (c.1880G>C, p.Arg627Thr, ENST00000343597) is absent from large population databases, falls at the same amino acid position (p.Arg627Gly) and at a neighboring amino acid position (p.Tyr621Asp, p.Thr625Asn, p.Ser628Asn, p.Ser628Arg) to missense variants reported in individuals with hereditary fibrosing poikiloderma with tendon contractures, myopathy, and pulmonary fibrosis (POIKTMP), suggesting that this position may be critical for protein function (41), and explains all elements of the patient's phenotype (MIM 615704).

This resulted in an ACMG/AMP classification of LP, with the following criteria applied: PM1, PM5 Supporting, PM2, and PP4.

In P6, a duo-sequenced family (proband and unaffected father), the Invitae Moon team ranked the causal variant at position three, following heterozygous variants in *GATAD2B* and *ADAR*. The *GATAD2B* missense variant (c.884C>T, p.Ala295Val, ENST00000368655) has *in silico* prediction leaning towards non-deleterious (REVEL 0.13 – BP4 Moderate) (40) and is present in large population databases (6/152,160 alleles in gnomAD v3, 9/264,690 alleles in TOPMed), arguing against a causal role in dominantly inherited infantile-onset disease (GAND syndrome, MIM 615074). The *ADAR* missense variant (c.983G>A, p.Arg328Gln, ENST00000647597) is a ClinVar VUS (ClinVar variation ID: 806225) reported in association with recessive Aicardi-Goutieres syndrome (MIM 615010). The proband is, however, missing a second variant in *ADAR*, and the identified heterozygous variant has *in silico* prediction leaning towards non-deleterious (REVEL 0.23 – BP4 Supporting) (40). In contrast, the heterozygous missense variant in *KCND2* in the answer key (c.1207C>G, p.Pro403Ala, ENST00000331113), this is presumed *de novo* given that it was not inherited from the father, explains the patient's phenotype of global developmental delay, hypotonia, and visual impairment (42), is predicted to be deleterious by *in silico* prediction (REVEL 0.84 – PP3 Moderate) (40) and is absent from large population databases. This resulted in an ACMG/AMP classification of LP, with the following criteria applied: PM2 Supporting, PP3 Moderate, PS3 Supporting, PS4 Supporting, PS2 Moderate.

In P7, a trio-sequenced family, the Invitae Moon team ranked the causal variant at position two, following compound heterozygous variants in *SYNE1*, a splice donor variant (c.11082+1G>A, ENST00000367255) with a high spliceAI score of 0.99 predicting donor loss and a missense variant (c.8308T>A, p.Phe2770Ile, ENST00000367255) reported in ClinVar as a VUS (ClinVar variation ID: 288416, no phenotype reported) with *in silico* prediction leaning towards non-deleterious (REVEL 0.07 – BP4 Moderate) (40). Recessive variants in *SYNE1* are reported to manifest in arthrogryposis multiplex congenita (MIM 618484) and spinocerebellar ataxia (MIM 610743) and are a weak phenotype match for the proband with a predominantly dysmorphic and neurological phenotype. In contrast, the *de novo* frameshift variant in *EHMT1* (c.1051del, p.Asp351ThrfsTer66, ENST00000460843) in the answer key, which has previously been reported in Kleefstra syndrome (MIM 610253), is absent from large population databases. This frameshift results in a premature stop codon 66 amino acids downstream predicting it to result in a truncated or absent protein, which is established disease mechanism for *EHMT1* in autosomal dominant Kleefstra syndrome. This resulted in an ACMG/AMP classification of P, with the following criteria applied: PVS1, PS2 Moderate, PM2 Supporting.

Finally, in P11, a proband-only family, the Invitae Moon team ranked the causal variant at position four, following heterozygous variants in *SYNE1*, *TTN*, and *POLR2A*, respectively. The *SYNE1* variant (c.226-2dup, ENST00000367255) is reported in ClinVar with conflicting interpretations of pathogenicity (ClinVar variation ID: 279936). Though *SYNE1* is phenotypically in-keeping with the proband's phenotype of muscle dystrophy, the spliceAI score is only 0.08 (predicting acceptor gain and likely resulting in an in-frame protein) and the variant is present in large population databases (17/152,056 alleles in gnomAD v3, 33/264,690 alleles in TOPMed), arguing against a causative role in this ultrarare dominantly inherited disease. The *TTN* missense variant (c.79867G>A, p.Glu26623Lys, ENST00000589042) is absent from large population databases, however, has *in silico* prediction of deleteriousness leaning towards non-deleterious (REVEL 0.18 – BP4 Moderate) (40). The presence of rare (≤1% population allele frequency) heterozygous *TTN* missense variants in 83,657/125,748 (66.5%) individuals in gnomAD v2 highlights how frequently these are encountered. Finally, a non-coding 3'UTR variant was prioritized in *POLR2A* (n.6341A>G, ENST00000617998), a gene primarily associated with neurodevelopmental abnormalities (MIM 618603), thereby not in-keeping with the proband's adult-onset, isolated muscular phenotype. In contrast, the causal missense variant in the answer key in *TPM2* (c.344A>T, p.Glu115Val, ENST00000645482), a gene associated with Nemaline myopathy (MIM 609285), is a phenotype match. The variant is absent from large population databases and has supporting *in silico* prediction (REVEL 0.89 – PP3 Moderate) (40), resulting in an ACMG/AMP classification of VUS, with the following criteria applied: PM2, PP3 Moderate. This variant had the least evidence of pathogenicity at present but was correctly prioritized in the top 5 variants by 40 of 52 models and was prioritized as the top candidate by 17 models.

### Supplemental Figures

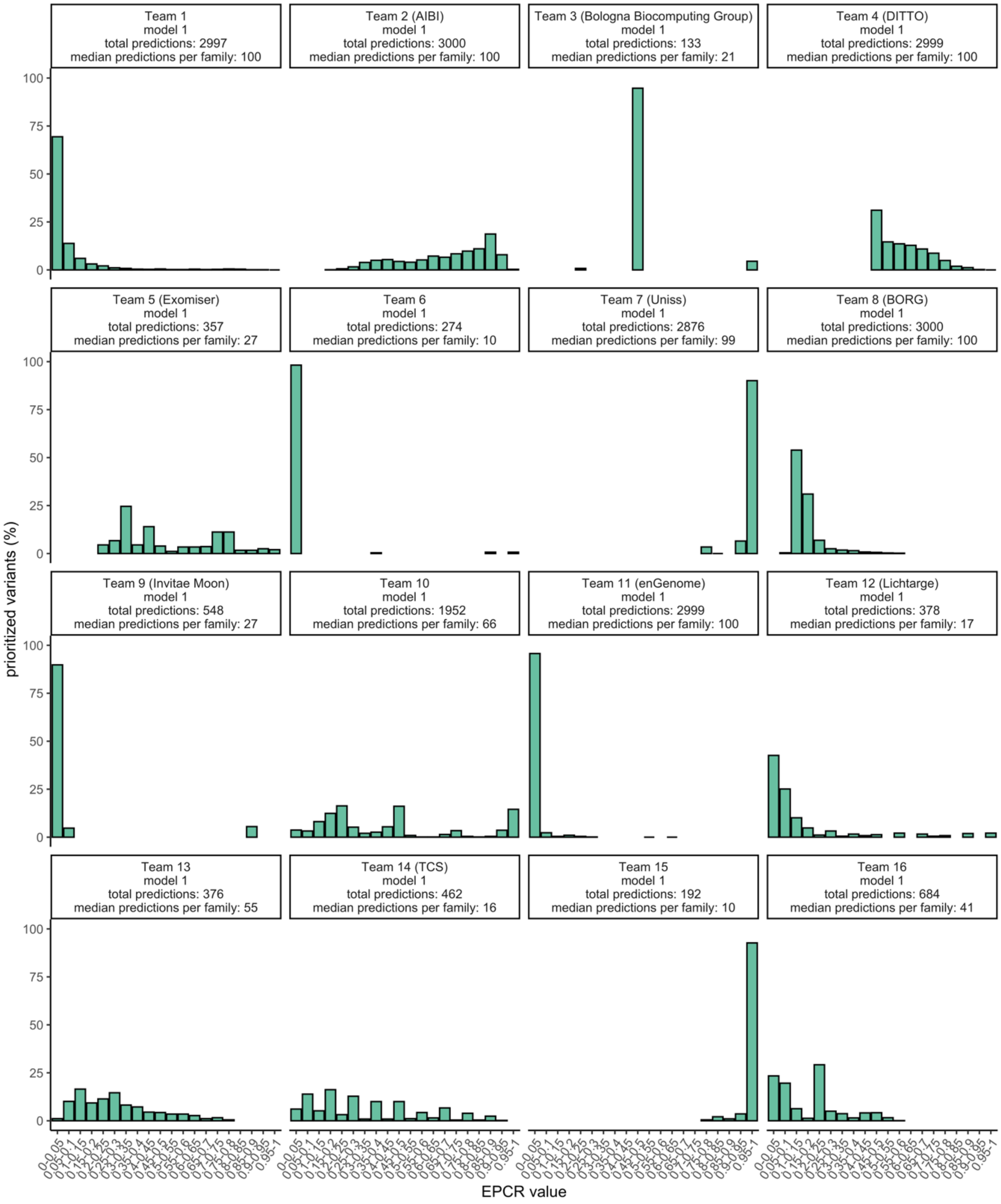


**Supplemental Figure 1.** Distribution of EPCR value for all variant predictions submitted by each team's primary model (model 1), indicating the total number of variant predictions submitted and the median number per family.

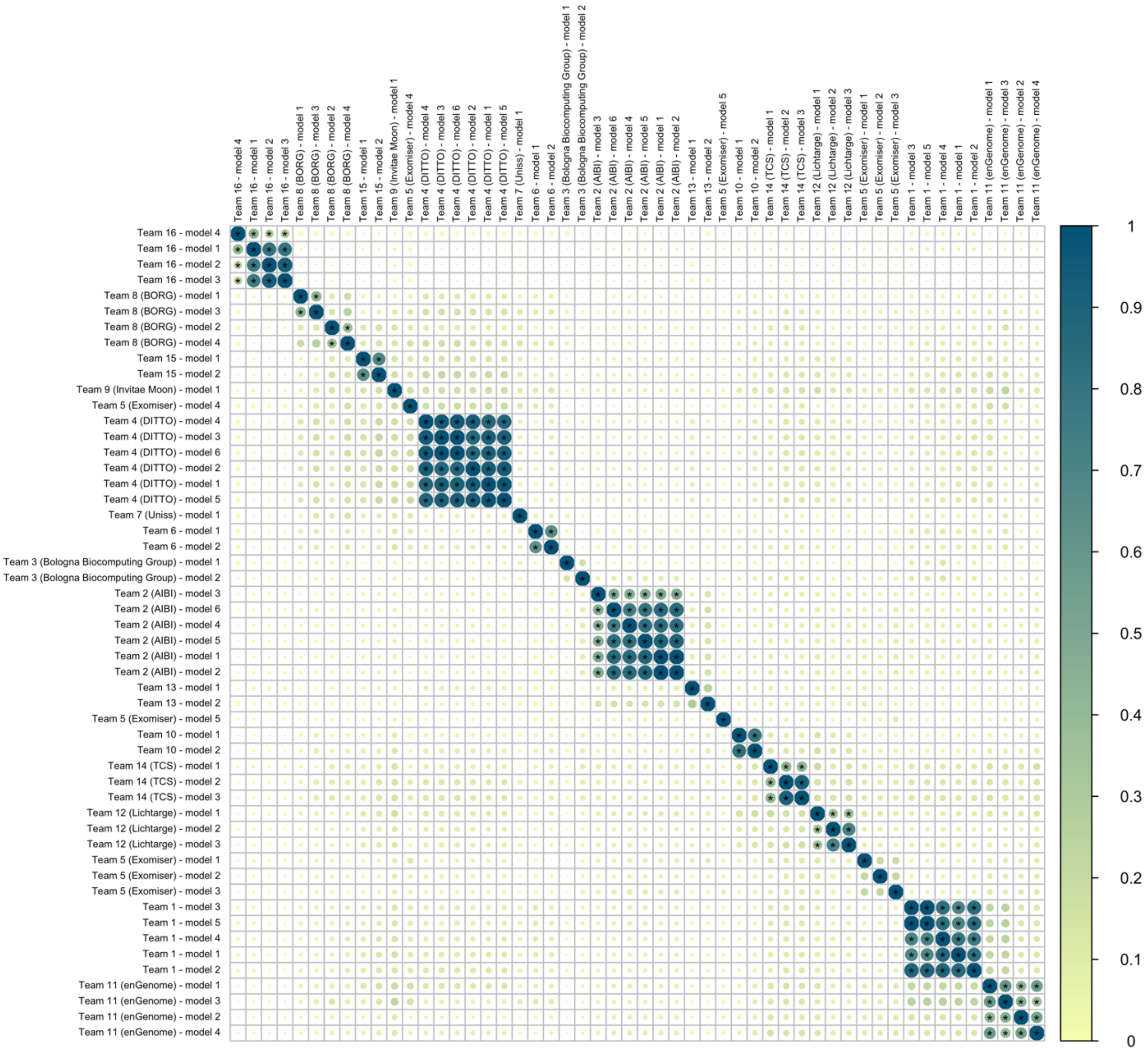


**Supplemental Figure 2.** Concordance between models for the top five ranked predictions per proband across all 30 families in the test set. Statistically significant values after Bonferroni correction for multiple testing are indicated with an asterisk. Missing predictions were considered discordant.

### Supplemental Tables

**Table S1. CAGI6 RGP challenge answer key for the 35 selected families in the training set** (see separate file)

**Table S2. CAGI6 RGP challenge answer key for the 30 selected families in the test set** (see separate file)

**Table S3. Returnable diagnoses and novel disease gene candidates in previously unsolved families in the test set** (see separate file)

### Supplemental References


1. Chen J. A fully-automated event-based variant prioritizing solution to the CAGI5 intellectual disability gene panel challenge. Hum Mutat. 2019 Sep;40(9):1364–72.

2. McLaren W, Gil L, Hunt SE, Riat HS, Ritchie GRS, Thormann A, et al. The Ensembl Variant Effect Predictor. Genome Biol. 2016 Jun 6;17(1):122.

3. Ioannidis NM, Rothstein JH, Pejaver V, Middha S, McDonnell SK, Baheti S, et al. REVEL: An Ensemble Method for Predicting the Pathogenicity of Rare Missense Variants. Am J Hum Genet. 2016 Oct 6;99(4):877–85.

4. Yang H, Robinson PN, Wang K. Phenolyzer: phenotype-based prioritization of candidate genes for human diseases. Nat Methods. 2015 Sep;12(9):841–3.

5. Lee J, Yoon W, Kim S, Kim D, Kim S, So CH, et al. BioBERT: a pre-trained biomedical language representation model for biomedical text mining. Bioinformatics. 2020 Feb 15;36(4):1234–40.

6. Babbi G, Martelli PL, Profiti G, Bovo S, Savojardo C, Casadio R. eDGAR: a database of Disease-Gene Associations with annotated Relationships among genes. BMC Genomics. 2017 Aug 11;18(Suppl 5):554.

7. Babbi G, Martelli PL, Casadio R. PhenPath: a tool for characterizing biological functions underlying different phenotypes. BMC Genomics. 2019 Jul 16;20(Suppl 8):548.

8. Bovo S, Martelli PL, Di Lena P, Casadio R. NETGE-PLUS: Standard and Network-Based Gene Enrichment Analysis in Human and Model Organisms. J Proteome Res. 2020 Jul 2;19(7):2873–8.

9. Greene D. ontologySimilarity [Internet]. 2021 [cited 2023 May 30]. Available from: https://cran.r-project.org/web/packages/ontologySimilarity/vignettes/ontologySimilarity-introduction.html

10. Orphanet [Internet]. [cited 2021 Dec]. Available from: https://www.orpha.net/consor/cgi-bin/index.php

11. Orphadata [Internet]. [cited 2021 Dec]. Available from: http://www.orphadata.org/cgi-bin/index.php

12. Narasimhan V, Danecek P, Scally A, Xue Y, Tyler-Smith C, Durbin R. BCFtools/RoH: a hidden Markov model approach for detecting autozygosity from next-generation sequencing data. Bioinformatics. 2016 Jun 1;32(11):1749–51.

13. Danecek P, Auton A, Abecasis G, Albers CA, Banks E, DePristo MA, et al. The variant call format and VCFtools. Bioinformatics. 2011 Aug 1;27(15):2156–8.

14. Wang K, Li M, Hakonarson H. ANNOVAR: functional annotation of genetic variants from high-throughput sequencing data. Nucleic Acids Res. 2010 Sep;38(16):e164.

15. Karczewski KJ, Weisburd B, Thomas B, Solomonson M, Ruderfer DM, Kavanagh D, et al. The ExAC browser: displaying reference data information from over 60 000 exomes. Nucleic Acids Res. 2017 Jan 4;45(D1):D840–5.

16. Collins RL, Brand H, Karczewski KJ, Zhao X, Alföldi J, Francioli LC, et al. A structural variation reference for medical and population genetics. Nature. 2020 May;581(7809):444–51.

17. McLaren W, Pritchard B, Rios D, Chen Y, Flicek P, Cunningham F. Deriving the consequences of genomic variants with the Ensembl API and SNP Effect Predictor. Bioinformatics. 2010 Aug 15;26(16):2069–70.

18. Rentzsch P, Witten D, Cooper GM, Shendure J, Kircher M. CADD: predicting the deleteriousness of variants throughout the human genome. Nucleic Acids Res. 2019 Jan 8;47(D1):D886–94.

19. Pedersen BS, Brown JM, Dashnow H, Wallace AD, Velinder M, Tristani-Firouzi M, et al. Effective variant filtering and expected candidate variant yield in studies of rare human disease. NPJ Genom Med. 2021 Jul 15;6(1):60.

20. Chen J, Althagafi A, Hoehndorf R. Predicting candidate genes from phenotypes, functions and anatomical site of expression. Bioinformatics. 2021 May 5;37(6):853–60.

21. Ashburner M, Ball CA, Blake JA, Botstein D, Butler H, Cherry JM, et al. Gene ontology: tool for the unification of biology. Nature genetics. 2000;25(1):25–9.

22. Smith CL, Eppig JT. The mammalian phenotype ontology: enabling robust annotation and comparative analysis. Wiley Interdiscip Rev Syst Biol Med. 2009 Nov-Dec;1(3):390–9.

23. Robinson PN, Köhler S, Bauer S, Seelow D, Horn D, Mundlos S. The Human Phenotype Ontology: a tool for annotating and analyzing human hereditary disease. Am J Hum Genet. 2008 Nov;83(5):610–5.

24. Rodríguez-García MÁ, Gkoutos GV, Schofield PN, Hoehndorf R. Integrating phenotype ontologies with PhenomeNET. J Biomed Semantics. 2017 Dec 19;8(1):58.

25. Amberger J, Bocchini C, Hamosh A. A new face and new challenges for Online Mendelian Inheritance in Man (OMIM). Human mutation. 2011;32(5):564–7.

26. Chen S, Francioli LC, Goodrich JK, Collins RL, Kanai M, Wang Q, et al. A genome-wide mutational constraint map quantified from variation in 76,156 human genomes [Internet]. bioRxiv. 2022 [cited 2023 May 30]. p. 2022.03.20.485034. Available from: https://www.biorxiv.org/content/10.1101/2022.03.20.485034v2

27. Sudlow C, Gallacher J, Allen N, Beral V, Burton P, Danesh J, et al. UK biobank: an open access resource for identifying the causes of a wide range of complex diseases of middle and old age. PLoS Med. 2015 Mar;12(3):e1001779.

28. Köhler S, Gargano M, Matentzoglu N, Carmody LC, Lewis-Smith D, Vasilevsky NA, et al. The Human Phenotype Ontology in 2021. Nucleic Acids Res. 2021 Jan 8;49(D1):D1207–17.

29. Piñero J, Ramírez-Anguita JM, Saüch-Pitarch J, Ronzano F, Centeno E, Sanz F, et al. The DisGeNET knowledge platform for disease genomics: 2019 update. Nucleic Acids Res. 2020 Jan 8;48(D1):D845–55.

30. Landrum MJ, Lee JM, Benson M, Brown GR, Chao C, Chitipiralla S, et al. ClinVar: improving access to variant interpretations and supporting evidence. Nucleic Acids Res. 2018 Jan 4;46(D1):D1062–7.

31. Stelzer G, Plaschkes I, Oz-Levi D, Alkelai A, Olender T, Zimmerman S, et al. VarElect: the phenotype-based variation prioritizer of the GeneCards Suite. BMC Genomics. 2016 Jun 23;17 Suppl 2(Suppl 2):444.

32. Jiang Y, Urresti J, Pagel KA, Pramod AB, Iakoucheva LM, Radivojac P. Prioritizing de novo autism risk variants with calibrated gene- and variant-scoring models. Hum Genet. 2022 Oct;141(10):1595–613.

33. Liu X, Jian X, Boerwinkle E. dbNSFP: a lightweight database of human nonsynonymous SNPs and their functional predictions. Hum Mutat. 2011 Aug;32(8):894–9.

34. Li B, Krishnan VG, Mort ME, Xin F, Kamati KK, Cooper DN, et al. Automated inference of molecular mechanisms of disease from amino acid substitutions. Bioinformatics. 2009 Nov 1;25(21):2744–50.

35. Szklarczyk D, Gable AL, Nastou KC, Lyon D, Kirsch R, Pyysalo S, et al. The STRING database in 2021: customizable protein-protein networks, and functional characterization of user-uploaded gene/measurement sets. Nucleic Acids Res. 2021 Jan 8;49(D1):D605–12.

36. Xin J, Mark A, Afrasiabi C, Tsueng G, Juchler M, Gopal N, et al. High-performance web services for querying gene and variant annotation. Genome Biol. 2016 May 6;17(1):91.

37. Karczewski KJ, Francioli LC, Tiao G, Cummings BB, Alföldi J, Wang Q, et al. The mutational constraint spectrum quantified from variation in 141,456 humans. Nature. 2020 May;581(7809):434–43.

38. Landrum MJ, Lee JM, Riley GR, Jang W, Rubinstein WS, Church DM, et al. ClinVar: public archive of relationships among sequence variation and human phenotype. Nucleic Acids Res. 2014 Jan;42(Database issue):D980-5.

39. Szklarczyk D, Franceschini A, Wyder S, Forslund K, Heller D, Huerta-Cepas J, et al. STRING v10: protein-protein interaction networks, integrated over the tree of life. Nucleic Acids Res. 2015 Jan;43(Database issue):D447-52.

40. Pejaver V, Byrne AB, Feng B-J, Pagel KA, Mooney SD, Karchin R, et al. Calibration of computational tools for missense variant pathogenicity classification and ClinGen recommendations for PP3/BP4 criteria. Am J Hum Genet. 2022 Dec 1;109(12):2163–77.

41. Mercier S, Küry S, Salort-Campana E, Magot A, Agbim U, Besnard T, et al. Expanding the clinical spectrum of hereditary fibrosing poikiloderma with tendon contractures, myopathy and pulmonary fibrosis due to FAM111B mutations. Orphanet J Rare Dis. 2015 Oct 15;10:135.

42. Zhang Y, Tachtsidis G, Schob C, Koko M, Hedrich UBS, Lerche H, et al. KCND2 variants associated with global developmental delay differentially impair Kv4.2 channel gating. Hum Mol Genet. 2021 Nov 16;30(23):2300–14.
